# Contractile cell apoptosis regulates airway smooth muscle remodeling in asthma

**DOI:** 10.1101/2021.02.08.21251269

**Authors:** Rebeca Fraga-Iriso, Óscar Amor-Carro, Nadia S. Brienza, Laura Núñez-Naveira, Beatriz Lema-Costa, Francisco Javier González-Barcala, Teresa Bazús-González, Carmen Montero-Martínez, Antolín López-Viña, Alfons Torrego, Vicente Plaza, Carlos Martínez-Rivera, Qutayba Hamid, James G. Martin, Patrick Berger, David Ramos-Barbón

**Author notes:** **Address correspondence to:** David Ramos-Barbón, MD, PhD. Respiratory Medicine Department, Mòdul 2-4^a^ Plta., Hospital de la Santa Creu i Sant Pau, Sant Antoni Maria Claret 167, 08025 Barcelona, Spain. **In memoriam** (Teresa Bazús-González). **Conflict of interest:** The authors have declared that no conflict of interest related to this work exists.

## Abstract

**Rationale:** Investigations on the mechanisms of airway smooth muscle remodeling, a prominent asthma feature contributing to its clinical manifestations and severity, have largely focused on its hyperplastic growth. Conversely, limited data and virtually no translational research have been produced on a plausible role of apoptosis in the homeostasis and remodeling of airway smooth muscle.

**Objectives:** We aimed at demonstrating an involvement of apoptosis, an essential regulator of organ structure and cell turnover, in the pathophysiology of airway smooth muscle remodeling in asthma.

**Methods:** Murine experimental asthma was modeled to analyze airway hyperresponsiveness, contractile tissue remodeling and apoptosis detection outcomes at early and late cutoffs, and under pharmacological inhibition of apoptosis by employing a caspase blocker. Clinical investigation followed through analyses on human bronchial biopsies.

**Results:** Airway hyperresponsiveness and contractile tissue remodeling were already established in early experimental asthma, and a subsequent upregulation of apoptosis limited the airway contractile tissue growth. Caspase inhibition elicited chaotic pulmonary mechanics and an unusual growth of airway smooth muscle that was structurally disorganized. In bronchial biopsies, airway smooth muscle increased from controls through subjects with intermittent and persistent moderate and severe asthma. Cleaved poly-ADP ribose polymerase (c-PARP, a byproduct of caspase activity) was increased in severe asthma.

**Conclusions:** Apoptosis is involved in airway contractile cell turnover and in shaping the size, structure and proper function of the airway smooth muscle layer. Apoptosis inhibitors may complicate concomitant asthma, whereas agents favouring airway contractile cell apoptosis may provide a novel pipeline of therapeutic development.

**Key messages:**

- How normal airway smooth muscle structure is preserved, and whether counteracting responses to remodeling are elicited in asthma, are outstanding questions not probed in vivo nor in the clinical setting.
- In this work, combined investigations on murine experimental asthma and human bronchial biopsies show that airway contractile cell apoptosis is involved in the homeostasis of airway smooth muscle, and apoptotic activity is upregulated as part of the remodeling process of this tissue in asthma.
- Apoptosis arises as a key regulator of the size and structure of the airway smooth muscle layer. This concept draws implications for clinical practice and drug development.

## Introduction

The evidence that airway smooth muscle remodeling is a prominent contributor to the pathophysiology and severity of asthma (1-11) has led to extensive investigations largely focused on the mechanisms of its hyperplastic growth (9, 11-20), whereas an almost absolute lack of data remains on any involvement of apoptosis, an essential regulator of morphogenesis and tissue homeostasis (21, 22). Early work where airway contractile tissue (AwCT) cell apoptosis was explored *in vivo* was a rat experimental asthma study showing a combined contribution of increased myocyte proliferation and inhibition of apoptosis, resulting from T cell driven AwCT remodeling (18). Contrary to such short-term rat model of induced disease, airway myocyte apoptosis was increased along with proliferation in horses with heaves, which represent spontaneous, long-term chronicity (23). Those previous studies led us to hypothesize that AwCT cell apoptosis is involved in the remodeling process and is subjected to regulation during the development of the disease. Here we aimed at providing proof-of-concept of an involvement of AwCT cell apoptosis in the pathophysiology of airway smooth muscle remodeling in asthma, by combining pharmacological inhibition of apoptosis in experimental disease modeling with clinical research through bronchial biopsies. Some preliminary results of these studies have been previously reported in the form of abstracts (24-27).

## Methods

A detailed methods section is issued in the online supplement.

### Experimental asthma

The murine experimental asthma studies were performed with institutional Animal Ethics Committee approval in compliance with European Union Directive 86/609 (28), and animal care and procedures were carried out as per subsequent guidelines detailed in Royal Decree 1201/2005 of Spain (29). Reporting followed the ARRIVE (Animal Research: Reporting of In Vivo Experiments) guidelines (30). BALB/c mice were sensitized to ovalbumin (OVA) and repeatedly challenged with OVA or phosphate-buffered saline (PBS) through intranasal (i.n.) instillation. Pulmonary resistance (R_L_) values in response to methacholine challenge were measured by forced oscillation technique with FlexiVent equipment. Bronchoalveolar lavage (BAL) was collected for total and differential leukocyte counts, and the lungs were inflated at standard pressure and fixed.

### Bronchial biopsies

Bronchial biopsies were collected in several centers as per the published guidelines (31), and subsequent recommendations (32), into a *Biobank Subspecialized in Bronchial Biopsy for Asthma Research*, a collection hosted as part of an institutional biobank at the Biomedical Research Institute of the *Hospital de la Santa Creu i Sant Pau, Universitat Autònoma de Barcelona*, with registration B.0000722 at the National Registry of Biobanks (*Instituto de Salud Carlos III*, Government of Spain), in compliance with Law 14/2007 on Biomedical Research (33). Bronchial biopsy collection for biobank hosting and investigational purposes was approved by an Institutional Review Board at all participating centers, and all subjects signed an Informed Consent Form. Bronchial biopsy sections from some of the subjects herein included were employed for other investigational goals and procedures in previous work (11). Biopsies were collected from control subjects, subjects with intermittent asthma managed in GINA-1 step, and subjects with persistent moderate and severe asthma.

### Biological specimen analyses

Murine lung and bronchial biopsy tissue sections were processed for terminal-deoxynucleotidyl-transferase-dUTP-nick-end-labeling (TUNEL), and immunohistochemical detection of active caspase-3 and cleaved poly-(ADP-ribose)-polymerase (c-PARP). The signals were colocalized with α-SMA co-immunostaining. Quantitative morphology was performed to measure the frequency of TUNEL^+^α-SMA^+^, active caspase-3^+^α-SMA^+^, and c-PARP^+^α-SMA^+^ cells. Airway contractile tissue (AwCT) mass, a dimensionless index, was measured from the α-SMA signal.

### Data analysis

Values are expressed as mean ± standard error of the mean (SEM) unless otherwise stated. Distributions were compared using Student’s t test for two-group comparisons where applicable, or one-way ANOVA otherwise. Post-ANOVA pairwise comparisons were analyzed with Fisher’s least significant difference test or Games-Howell test for unequal variances as appropriate. One-tailed Dunnett’s test versus single control was employed in the case of intra-group R_L_ comparisons versus baseline R_L_. A *P* value of less than 0.05 was considered statistically significant. The effect size was estimated by the 95% confidence interval (CI) of the mean increment and expressed as (mean Δ [CI]). SPSS version 19 (SPSS Inc., IBM Corporation, New York, US) was employed for statistical analysis (34).

## Results

Numerical data and *P* values are provided in the online supplement.

### Airway α-SMA^+^ cell apoptosis is upregulated subsequent to early development of airway hyperresponsiveness (AHR) and increased AwCT mass

We first profiled the frequency of AwCT cell apoptosis in murine experimental asthma models representing early and long-term stages of disease development and severity. OVA-sensitized mice were i.n.-challenged with OVA (OVA group) or PBS as control (CTRL group), 3 times weekly for 1 week in the early-disease model and 12 weeks in the long-term model (n=8 for all groups; design in Figure 1-A). At the early-disease cutoff, the OVA group showed already established AHR upon methacholine challenge at 48 h after the last i.n. instillation, as demonstrated by a significant increase of pulmonary resistance (R_L_) over intra-group baseline R_L_ and also versus the CTRL group, with a maximal mean R_L_ increment of 2.2-fold over CTRL (Figure 1B; numerical data and *P* values in supplemental Table E1). As a quantitative AwCT remodeling outcome we analyzed the α-SMA^+^ section surface area referenced to airway basement membrane perimeter squared (P_BM_^2^) (18, 35), a dimensionless variable termed AwCT mass to accurately reflect that α-SMA is expressed by airway smooth muscle cells and myofibroblasts (6). AwCT was significantly increased in the OVA animals at both the early-disease (4.28-fold versus CTRL) and long-term (2.56-fold) cutoffs. In the early-disease OVA animals, the AwCT increment was also higher (1.28-fold) than in their counterpart long-term cutoff animals (Figure 1C and supplemental Table E2). The data demonstrated therefore the presence of AHR and established AwCT remodeling at the early-disease cutoff. The increase in AwCT mass at such an early disease stage, which then plateaued to show attenuation at the long-term cutoff, suggested that α-SMA^+^ cell apoptosis might play a role in limiting AwCT overgrowth as the experimental disease progresses. To probe this idea, we performed colocalization of TUNEL with α-SMA. Alpha-SMA^+^TUNEL^+^ cells were examined for the identification of AwCT cell apoptosis, and quantified and corrected for airway size by dividing by P_BM_^2^. Active caspase-3, the final effector caspase in the programmed cell death pathway, was also co-immunostained with α-SMA for further supportive evidence of apoptosis. In the early-disease cutoff, no AwCT cell apoptosis was found in the OVA animals over the CTRL baseline. In the long-term cutoff, the OVA animals showed increased numbers of α-SMA^+^TUNEL^+^ (5.98-fold versus CTRL) and α-SMA^+^/active caspase-3^+^ (3.91-fold) cells/mm^2^ (Figure 1 D-E and Table E3). AwCT cell apoptosis was therefore significantly increased in the OVA animals at the long-term cutoff, by 7.18-fold over early-disease OVA animals as per α-SMA/TUNEL co-localization and 6.25-fold as per α-SMA/active caspase-3. The data suggested that, following the early presence of AwCT remodeling and AHR, apoptosis is upregulated in response to AwCT overgrowth in the course of experimental asthma.

**Figure 1.**
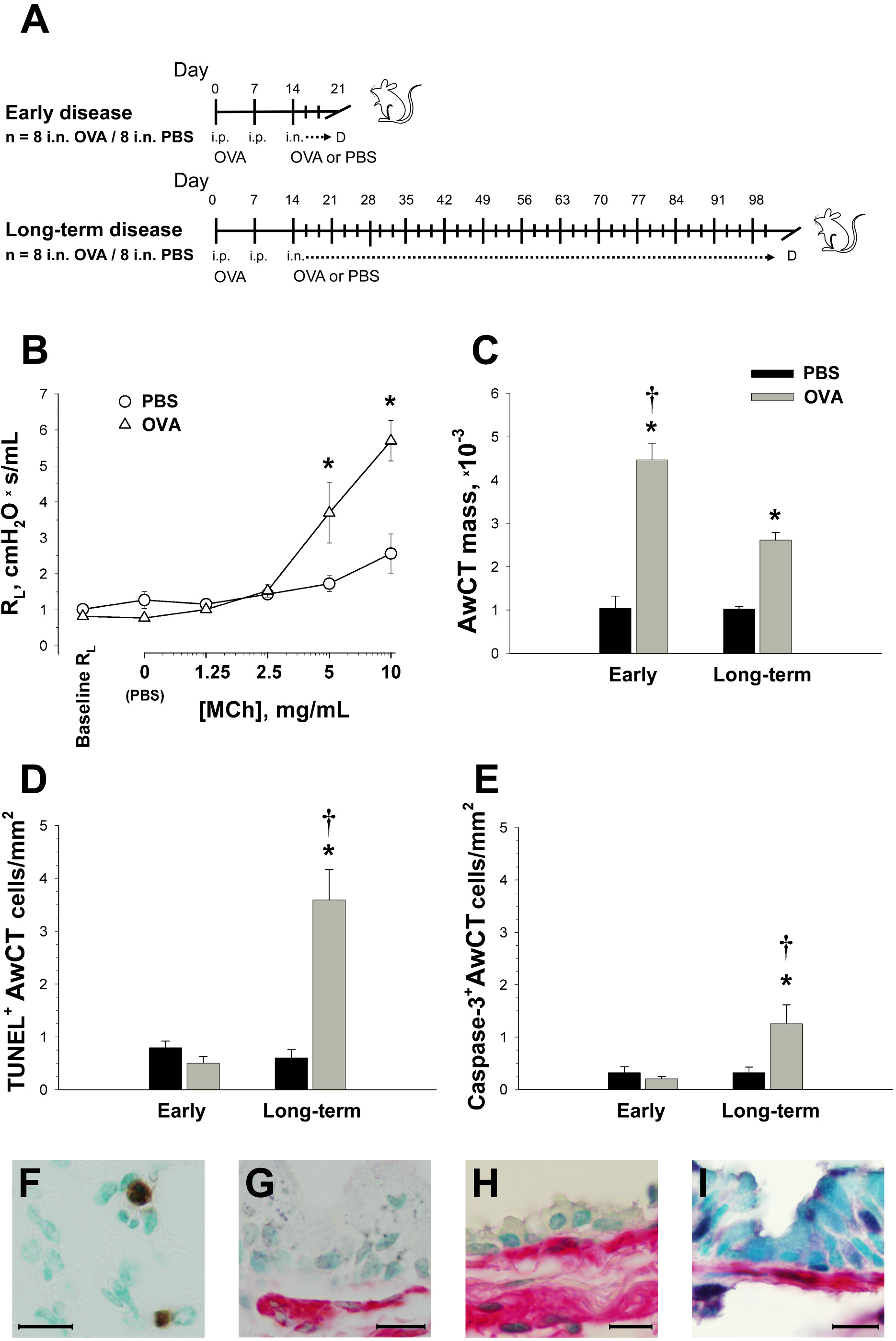
Airway responsiveness and AwCT apoptosis in early and long-term experimental asthma. (**A**) Mice were OVA-sensitized and i.n. challenged with OVA or PBS (control group, CTRL), 3 times weekly for 1 or 12 weeks. (**B**) Airway responsiveness to methacholine (MCh) at the early disease cutoff. (**C**) AwCT mass. (**D-E**) Assessment of AwCT apoptosis by co-localization of α-SMA with TUNEL and activated caspase-3, respectively. (**F-H**) Co-localization of α-SMA (red signal) with TUNEL (dark nuclear signal). (F) shows TUNEL^+^ inflammatory cells and (G-H) show airway wall sections from OVA-instilled animals, early and long-term disease respectively. (**I**) Co-localization of α-SMA with activated caspase-3 (dark nuclear signal), OVA-instilled animal at the long-term disease cutoff. Micrographs in (G-I) are oriented to show the airway epithelium on top. Airway hyperresponsiveness and increased AwCT mass were present at the early disease stage, and AwCT cell apoptosis was subsequently upregulated. *: *P*<0.05 versus PBS; †: *P*<0.05 versus early-disease OVA group. “D”: data acquisition cutoff in (A) diagram. Bar legend in (C) applies to (C-E). Scale bars: 20 µm.

### Pharmacological inhibition of apoptosis through experimental asthma elicits chaotic pulmonary mechanics and unusual, disorganized AwCT growth

To provide experimental evidence on the upregulation of apoptosis as a compensatory mechanism for the hyperplastic AwCT growth in airway remodeling, we modeled asthma while inhibiting apoptosis using the broad-spectrum caspase inhibitor QVD-OPH (36, 37). Mice were OVA sensitized and airway challenged with OVA or PBS, and each experimental arm was dosed i.p. with QVD-OPH or DMSO vehicle throughout the i.n. instillation period. A data collection cutoff point was chosen as the mid-point between the early and long-term disease models, i.e. at 21 i.n. instillations (experimental design in Figure 2A). OVA-challenged mice that received DMSO ([i.n. OVA + i.p. DMSO] group) showed AHR to methacholine as for usual experimental asthma, compared with the [i.n. PBS + i.p. DMSO] and [i.n. PBS + i.p. QVD-OPH] control groups (Figure 2 and supplemental Table E4). Conversely, mice that were OVA challenged under treatment with QVD-OPH ([i.n. OVA + i.p. QVD-OPH] group) showed a distinct and unusual airway response to methacholine challenge. This consisted of a highly variable R_L_ combining features such as an elevated baseline value, increases at low methacholine doses, sharp peak-and-valley sequences, poor correlation between methacholine dose and response, and dissociation between the R_L_ and elastance tracings (Figure 2).

**Figure 2.**
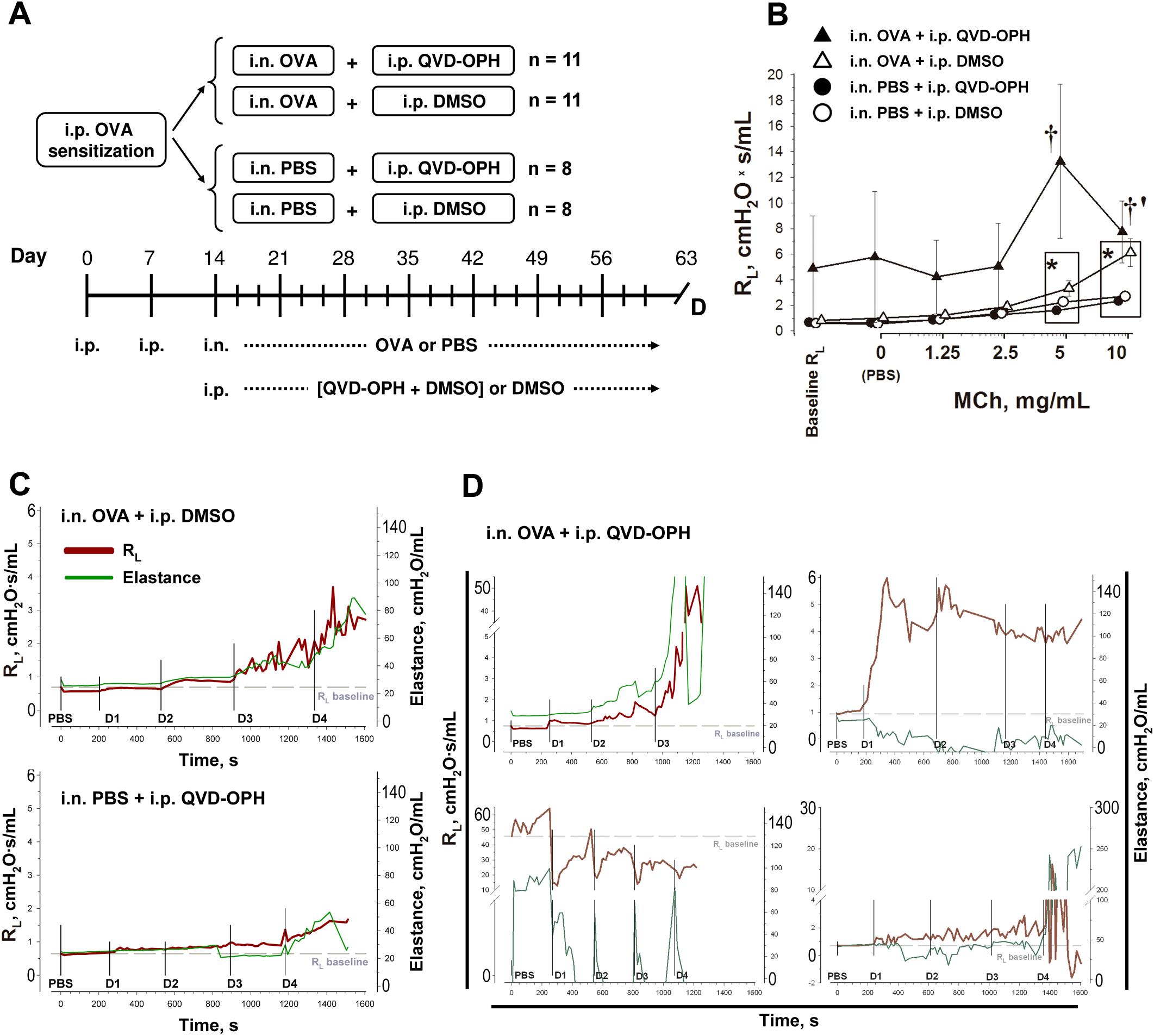
Airway methacholine challenge responses under pharmacological inhibition of apoptosis. (**A**) Mice were i.p. OVA-sensitized and i.n. instilled with OVA or PBS, 3 times weekly for 7 weeks. Intranasal instillations were accompanied by i.p. administration of QVD-OPH or DMSO vehicle. “D”: data acquisition cutoff. (**B**) Airway responsiveness to methacholine (MCh). **\***: *P*<0.05 versus intra-group baseline R_L_. **†**: *P*<0.05 versus [i.n. PBS + i.p. QVD-OPH] and [i.n. PBS + i.p. DMSO]. **‡**: *P*<0.05 versus all other groups. **§**: n = 9 for the [i.n. OVA + i.p. QVD-OPH] group at this data point due to the death of two animals before the 10 mg/mL MCh administration. (**C-D**) Illustrative examples of individual R_L_ and elastance tracing from [i.n. OVA + i.p. DMSO], [i.n. PBS + i.p. QVD-OPH] and [i.n. OVA + i.p. QVD-OPH] animals as indicated. The plots in (D) represent patterns of unusual airway responses to MCh challenge including exaggerated R_L_ and elastance increases through the progression of MCh concentrations, early steep R_L_ rises, and “needle” sequences starting from lifted or normal baseline values. Abscissae were normalized to 0-6 cmH_2_O·s/mL R_L_ and 0-140 cmH_2_O/mL elastance. Axis breaks were used where necessary to represent animals exceeding those value ranges.

On microscopic examination, the [i.n. OVA + i.p. QVD-OPH] animals showed an unusual pattern of airway smooth muscle remodeling, as compared with the untreated [i.n. OVA + i.p. DMSO] group. The pattern of remodeling consisted of increased thickening of the airway smooth muscle layer along with morphological features of disorganized growth (Figure 3). The quantitative analysis of AwCT on α-SMA immunostained specimens revealed a 1.7-fold increase in the [i.n. PBS + i.p. QVD-OPH] group over the [i.n. PBS + i.p. DMSO] animals, attributable to altered baseline AwCT homeostasis under the effect of QVD-OPH, and a 1.6-fold excess growth in the [i.n. OVA + i.p. QVD-OPH] animals over the [i.n. OVA + i.p. DMSO] animals (Figure 3 and supplemental Table E5).

**Figure 3.**
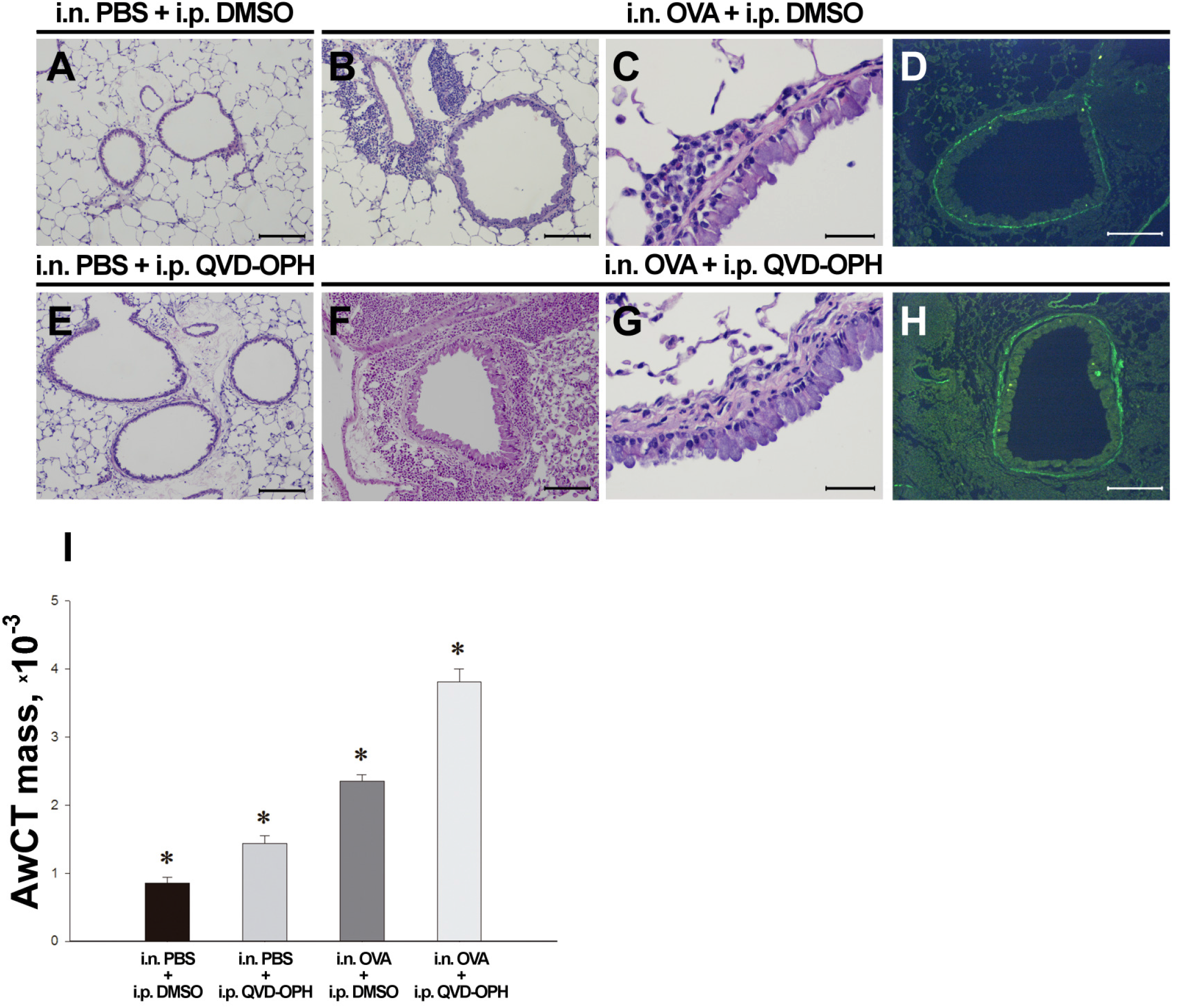
Effect of pharmacological inhibition of apoptosis on AwCT remodeling. (**A-H**) Micrographs representing the experimental arms as indicated. Lung sections were H&E stained as in (A-C) and (E-G), or α-SMA immunostained (D, H) for AwCT mass quantification. The high-magnification images in (C) and (G) allow for the appreciation of airway wall histopathology in usual experimental asthma ([i.n. OVA + i.p. DMSO], C) and the effect of running the experimental disease model under the inhibition of apoptosis by the QVD-OPH caspase blocker ([i.n. OVA + i.p. QVD-OPH], G). Usual airway remodeling features such as a thickened, hyperplastic epithelium and an increased airway smooth muscle layer can be appreciated in (C), accompanied by mononuclear and eosinophilic airway inflammation. In (G), a highly hyperplastic thickening of the airway smooth muscle layer is prominent. The layer morphology, atypical, is characterized by undulating smooth muscle fascicles with their nuclei laid out such as in a zig-zag pattern, and a loss of compactness reflected by the presence of interfascicular clefts. It was also noticeable that the [i.n. OVA + i.p. QVD-OPH] animals had profuse lung inflammation, such as shown in (F). (**I**) Quantification of AwCT mass. QVD-OPH generated a degree of AwCT increment in resting (i.n. PBS) animals, and potentiated the increase of AwCT mass in experimental asthma (i.n. OVA). Scale bars: 200 µm in (A-B), (D-F) and (H); 25 µm in (C, G). *: *P*<0.05 versus all other groups.

We investigated the effect of QVD-OPH on the frequency of AwCT cell apoptosis by co-localizing TUNEL and active caspase-3 with α-SMA (Figure 4 A-B, D-E, G and supplemental Table E6). The [i.n. OVA + i.p. DMSO] animals had their α-SMA^+^TUNEL^+^ cells increased by 4.46-fold over their [i.n. PBS + i.p. DMSO] counterpart group, a data set consistent with the upregulation of apoptosis seen in the prior exploratory experiments on early versus long-term disease modeling. The α-SMA^+^active caspase-3^+^ cell count showed a consistent trend but did not reach statistical significance. Conversely, both the α-SMA^+^TUNEL^+^ and α-SMA^+^active caspase-3^+^ cell counts were significantly decreased in the [i.n. OVA + i.p. QVD-OPH] versus the [i.n. OVA + i.p. DMSO] animals (−25.9% for α-SMA^+^TUNEL^+^ cells and −47.6% for α-SMA^+^active caspase-3^+^ cells), confirming an *in vivo* inhibitory effect of QVD-OPH on the AwCT cell apoptosis induced by experimental asthma. For additional verification that QVD-OPH downregulated apoptosis *in vivo* through caspase inhibition, we evaluated the activity of caspase-3 by analyzing the fragmentation of poly-(ADP-ribose)-polymerase (PARP), the main nuclear caspase-3 substrate (Figure 4 C, F, H and supplemental Table E6). For this purpose, we performed immunohistochemical detection of a 24kDa PARP fragment (cleaved PARP or c-PARP) generated from proteolytic cleavage by caspase-3, and co-localized this signal with α-SMA. QVD-OPH reduced the numbers of α-SMA^+^c-PARP^+^ cells in treated animals, whether PBS or OVA challenged, compared with the animals that received DMSO (by −33.7% for [i.n. PBS + i.p. QVD-OPH] versus [i.n. PBS + i.p. DMSO], and a borderline significant −28.3% for [i.n. OVA + i.p. QVD-OPH] versus [i.n. OVA + i.p. DMSO] animals).

**Figure 4.**
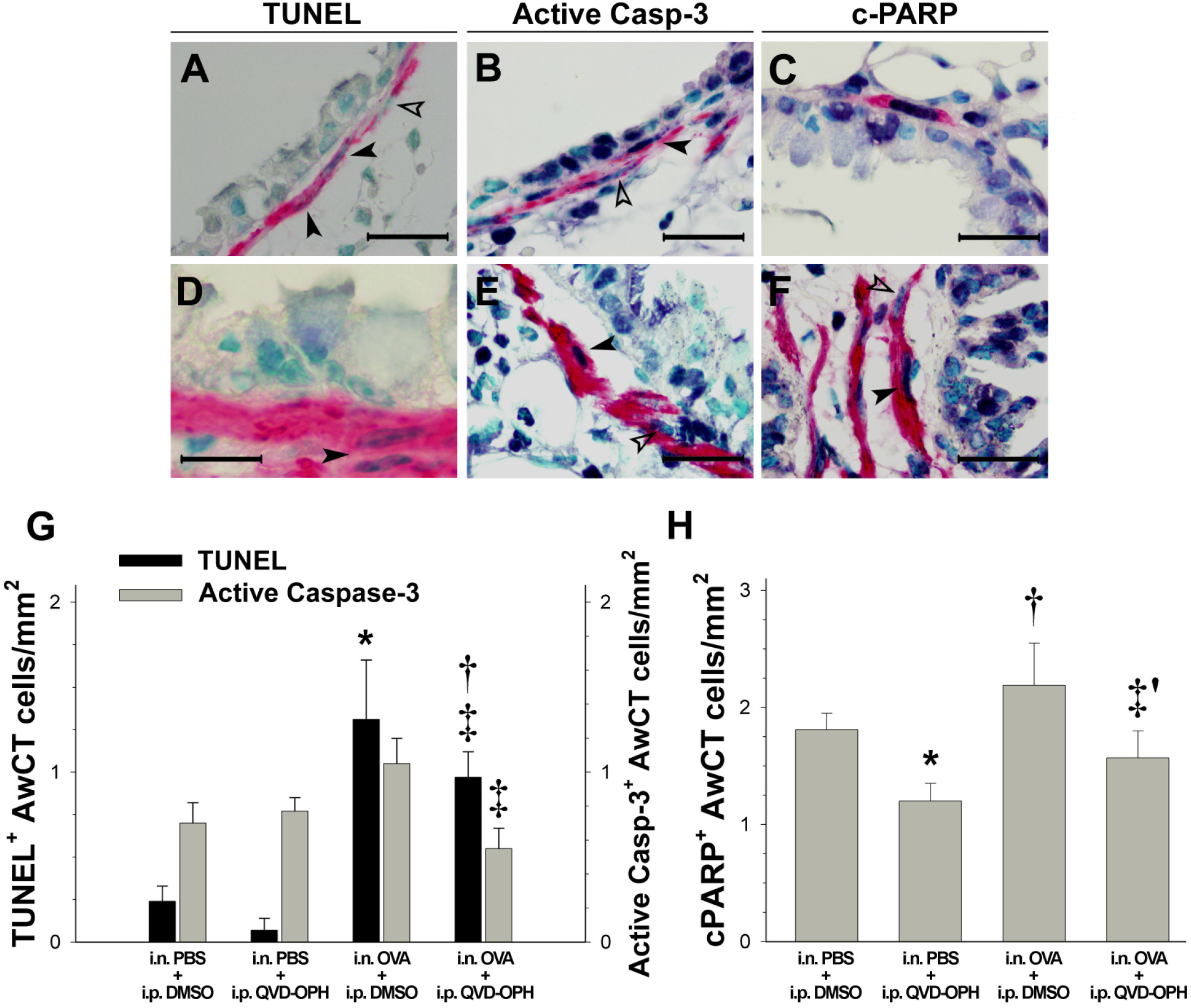
Effect of QVD-OPH on AwCT cell apoptotis *in vivo*. Colocalization of TUNEL (**A, D**), active caspase-3 (**B, E**) and c-PARP (**C, F**) with α-SMA. The micrographs represent [i.n. OVA + i.p. DMSO] (A-C) and [i.n. OVA + i.p. QVD-OPH] animals (D-F). Filled arrowheads signal examples of α-SMA^+^TUNEL^+^, α-SMA^+^active-caspase-3^+^ and α-SMA^+^c-PARP^+^ cells as indicated, and open arrowheads cells point to negative nuclei for reference. (**G-H**) Frequency of AwCT TUNEL^+^, active caspase-3^+^ and c-PARP^+^ cells, as indicated. Scale bars: 200 µm in (A-B, E); 100 µm in (D); 25 µm (C, F). *: *P*<0.05 versus [i.n. PBS + i.p. DMSO]; **†**: *P*<0.05 versus [i.n. PBS + i.p. QVD-OPH]; **‡**: *P*<0.05 versus [i.n. OVA + i.p. DMSO]; **‡’**: *P*=0,084.

The [i.n. OVA + i.p. DMSO] animals showed eosinophilic and lymphocytic airway inflammation in comparison with the [i.n. PBS + i.p. QVD-OPH] and [i.n. PBS + i.p. DMSO] control groups. QVD-OPH treatment did not lead to enhanced airway inflammation in the [i.n. OVA + i.p. QVD-OPH] group, as per total and differential leukocyte counts in BAL (see supplemental Figure E1 and Table E7).

### Airway smooth muscle cell apoptosis is present in human asthma and c-PARP release correlates with asthma severity

Our experimental data support that α-SMA^+^ cell apoptosis is involved in AwCT turnover and is further implicated in regulating the morphogenesis of airway smooth muscle under pathological conditions so as during the progression of murine experimental asthma. To search for evidence of such involvement of apoptosis in the pathophysiology of asthma, we analyzed bronchial biopsy sections from 59 subjects with asthma comprising 7 intermittent, 33 moderate and 19 severe asthmatics, plus 13 non-asthmatic control subjects. TUNEL co-localization with α-SMA immunostaining and quantitative morphology were performed to determine airway smooth muscle mass and the frequency of TUNEL^+^α-SMA^+^ cells (Figure 5A-F, J). Active caspase-3 immunostaining was not technically feasible in the human specimens. c-PARP was co-immunostained with α-SMA in the control subjects and in a sample subset of 5 moderate and 8 severe asthmatics (Figure 5G-I, K). Demographics and clinical data are provided in supplemental Table E8, and quantitative morphology data in Table E9.

**Figure 5.**
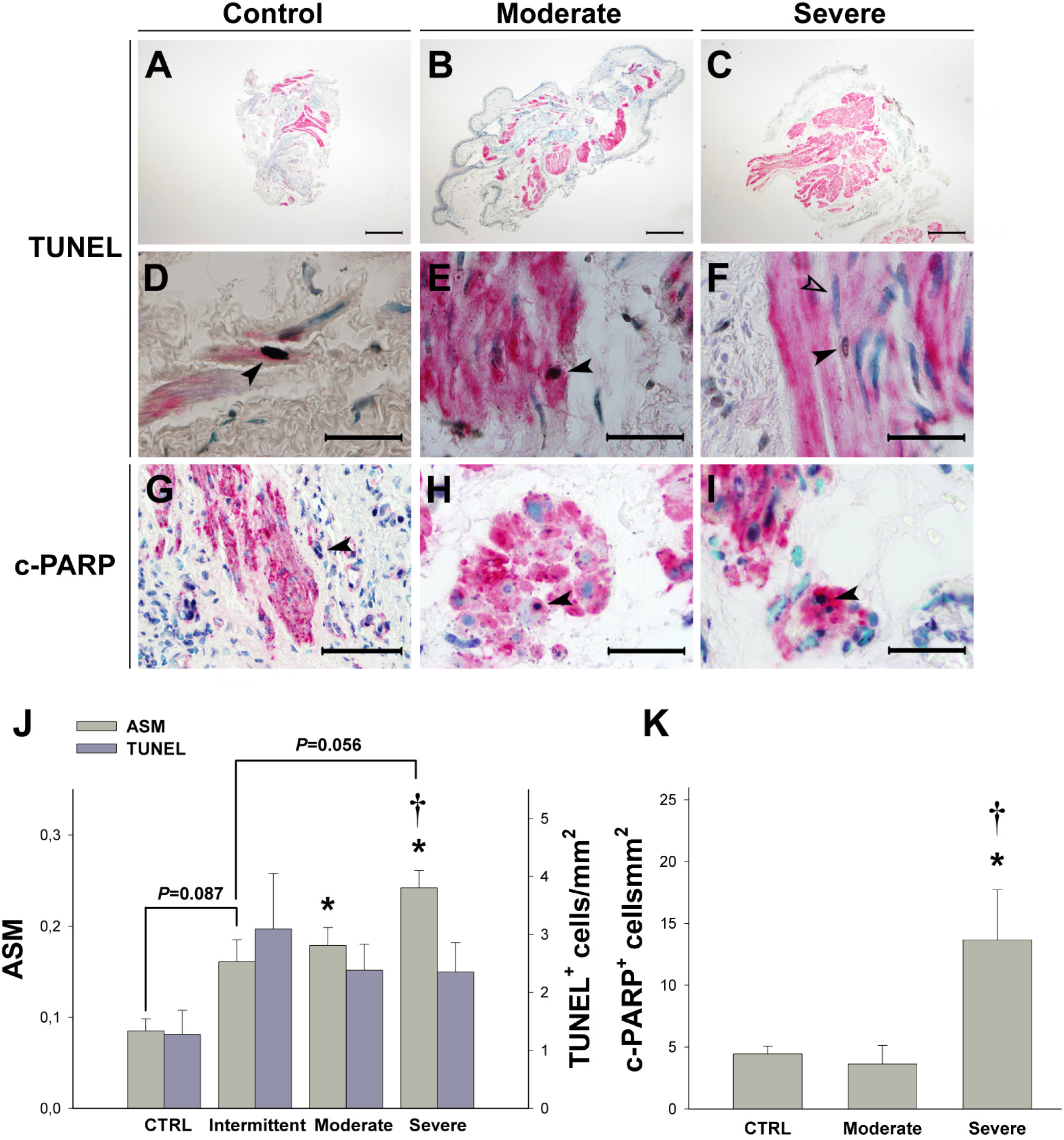
Apoptosis in human airway smooth muscle. Bronchial biopsy sections were processed for TUNEL (**A**-**F**), or immunostained for c-PARP detection (**G**-**I**), with signal co-localization with α-SMA. Biopsy sections from control subjects, moderate and severe asthmatics are shown as indicated. The low-magnification micrographs in (A-C) illustrate the airway smooth muscle progression from controls through moderate and severe asthmatics. In (D-F), examples of TUNEL^+^ myocyte nuclei (filled arrowheads) and non-apoptotic myocytes as an intra-specimen negative reference (open arrowhead) are shown. In (G-I), an increased frequency of α-SMA^+^c-PARP^+^ (dark nuclear signal) cells can be appreciated in the severe asthmatics. (**J**) Bronchial smooth muscle mass (ASM) and frequency of α-SMA^+^TUNEL^+^ cells. (**K**) Frequency of α-SMA^+^c-PARP^+^ cells. *: *P*<0.05 versus control subjects. †: *P*<0.05 versus moderate asthmatics. Borderline significant differences are specified. Scale bars: 300 µm (A-C); 50 µm (G); 20 µm (D-F, H-I).

Airway smooth muscle mass showed a significant gradient from the control subjects through the subjects with severe asthma. A tendency to increased airway smooth muscle was already present in the subjects with intermittent asthma versus controls (1.9-fold increment), with borderline statistical significance (*P*=0.087). Moderate asthmatics had significantly increased airway smooth muscle (2.1-fold) versus controls, and severe asthmatics showed a significant increase versus both controls (2.8-fold) and moderate asthmatics (1.3-fold). An observed greater frequency of TUNEL^+^α-SMA^+^ cells in the asthmatics did not yield a statistically significant result. However, severe asthmatics did show a significant increase in the frequency of c-PARP^+^α-SMA^+^ cells versus controls (3.1-fold) and moderate asthmatics (3.8-fold).

## Discussion

Despite apoptosis being a basic mechanism of life crucially involved in organ morphogenesis and tissue homeostasis (21, 22), virtually no attention has been paid to a potential role of apoptosis in airway smooth muscle remodeling, nor its plausible involvement in the cell turnover of this structure *in vivo*. Data on the expression of apoptosis related receptors by airway smooth muscle cells and their ability to undergo apoptosis are from few studies (38-42) chiefly based on cell culture, and no translational investigations have followed. As for *in vivo* data, the mismatch between short-term experimental asthma (18) and heaves (23) suggested that, should mechanisms leading to airway smooth muscle hyperplasia be activated at early asthma stages, some regulatory events would have to take place beyond a certain threshold of disease and remodeling, because the growth of airway smooth muscle in asthma is likely not unlimited. This rationale led us to conduct the combined murine experimental asthma and clinical studies presented here.

We first probed the frequency of AwCT cell apoptosis in murine experimental asthma at early and long-term disease cutoffs. For this purpose we co-localized α-SMA with TUNEL which, where applied for the *in situ* detection of apoptosis on tissue sections, yields high specificity if tissue necrosis, not a feature pertaining to asthma, is ruled out through proper histopathological scrutiny (43). Since AwCT cell apoptosis was the primary outcome, we wished to provide further supportive evidence by also co-localizing α-SMA with active caspase-3, a final effector caspase common to the receptor-mediated and mitochondrial apoptosis pathways (44). AHR and increased AwCT were already present at the early disease cutoff, a finding in line with some clinical observations showing that airway smooth muscle remodeling may develop in early stages of asthma (45-47). The low frequency of apoptotic events at such short-term cutoff may reflect baseline cell turnover for AwCT homeostasis. At the long-term disease cutoff, no further AwCT mass increment occurred, whereas the numbers of AwCT apoptotic cells were largely increased, consistently as per both TUNEL and active caspase-3 readouts. The data suggested that, following early AwCT remodeling and the onset of AHR, AwCT cell apoptosis is upregulated as a compensatory mechanism limiting AwCT growth.

Aiming at mechanistic demonstration of the role of apoptosis as a regulator of AwCT remodeling, we ran a new experimental asthma model while treating with the pan-caspase inhibitor QVD-OPH, a non-toxic drug that allows for effective pharmacological inhibition of apoptosis *in vivo* (36, 37). The OVA-challenged animals that received drug vehicle showed usual experimental asthma features, whereas the QVD-OPH treatment distorted the experimental disease and elicited unusual outcomes, with prominent and atypically patterned AwCT growth as a result. There was prominent airway wall thickening that was found to mostly occur at the expense of the airway smooth muscle layer which, beyond its enlargement over that of regular experimental asthma, showed morphological signs of tissue disorganization consisting of long, undulate fascicles with loss of bundle compaction. Such anomalous AwCT growth was associated with an unusual pattern of pulmonary mechanics and airway responsiveness to methacholine challenge. The animals posed difficult-to-manage mechanical ventilation and had aberrant responses to methacholine characterized by instability and unpredictability. It seems plausible that such chaotic pulmonary function resulted from “fibrillation” of an airway smooth muscle layer that was largely increased in its thickness, yet disorganized in its structure. Underlining such findings there was a limitation to AwCT cell apoptosis, resulting from the blocking effect of QVD-OPH, which reduced the increment in AwCT apoptosis shown by the experimental asthma animals treated with vehicle. The data therefore support the hypothesis that AwCT apoptosis is involved in the homeostasis of airway smooth muscle mass and, when the equilibrium of this tissue structure is challenged by remodeling as part of asthma pathophysiology, apoptosis turns into a counterbalancing regulator to limit airway smooth muscle growth, in addition to the preservation of its structure and function.

In view of the experimental asthma outcomes, the next aim was to seek for evidence of such involvement of AwCT cell apoptosis in human asthma. Thus, we analyzed bronchial biopsies representing a stepwise scale of asthma severity from control subjects, through subjects with intermittent asthma in GINA-1, and up to subjects with persistent, moderate and severe asthma. Quantitative morphology on α-SMA immunostained biopsy sections revealed a progression of airway smooth muscle mass in association with asthma severity, consistent with previously published data (11, 48, 49). An interesting novelty in the present study was the inclusion of a subset of adult subjects with intermittent asthma managed in therapeutic step GINA-1. Remarkably, airway smooth muscle remodeling was present in these subjects, whose increment in airway smooth muscle mass over controls was close to that of subjects with persistent, moderate asthma. The intermittent asthma versus controls achieving only borderline significance resulted from the study being underpowered within a 4-arm multiple comparison, and there was no difference between intermittent and persistent, moderate asthma. Although our study did not comprise age-guided sampling with inclusion of pediatric asthma (45-47), and despite that the natural history of asthma does not necessarily involve clinical progression from intermittent asthma through the severity steps of persistent asthma, the finding of airway smooth muscle remodeling in intermittent asthma appears consistent with the same finding at the early cutoff in our sequential experimental model. As for the evaluation of airway smooth muscle cell apoptosis, limited for technical reasons to TUNEL co-localization with α-SMA, a baseline frequency of myocyte apoptosis in the control subjects was present, suggesting again participation in homeostatic cell turnover. The observation of a trend towards overall greater numbers of TUNEL^+^ airway smooth muscle cells in the asthmatics neither yielded statistical significance versus controls, nor reflected any gradation associated with the sequence of airway smooth muscle increments in the asthmatics. However, the *in situ* analysis of the frequency of apoptotic cells may have limited the sensitivity for outcome detection in this setting. Apoptotic events at DNA nicking stage, the target of TUNEL detection, occur within a short time window and in a quite spatially scattered fashion (50). Contrary to the murine experimental models, where entire lung slices are examined, human bronchial biopsies are in comparison extremely minute samples of the airway tree. This likely compromises the suitability of bronchial biopsy as a sensitive procedure for the detection of significant outcomes from events that naturally occur with very low frequencies, such as is the case for TUNEL detection. Seeking an alternative approach we proceeded with co-localized detection of c-PARP as a byproduct of caspase-3 activity, which was demonstrably increased in severe asthma. This output was sound in terms of size of the difference and is consistent with a status of increased pro-apoptotic activity in the airway smooth muscle of the severe asthmatics.

In summary, the data presented here jointly provide conceptual support for a role of apoptosis in AwCT cell turnover, and in the regulation of the size and proper structure and function of the airway smooth muscle layer. The pharmaceutical industry has produced apoptosis inducer and inhibitor drugs that are either licensed or under clinical development for a variety of indications, from cancer to regenerative medicine. One straightforward implication of our findings for clinical practice is that asthmatic subjects, if treated or enrolled in trials with an apoptosis inhibitor for a concomitant condition, may need close monitoring of their airway structural integrity and function. Therapeutic development of apoptosis inducers for severe asthma, agents mostly confined as antineoplastic drugs at present, may pertain to a more complex and distant horizon. However, the data on bronchial thermoplasty (10) as the only existing therapy aimed at directly reducing the airway smooth muscle layer, contribute further support to a rationale for targeting airway smooth muscle cell apoptosis as a novel pipeline of development.

## Supporting information

ARRIVE Checklist

COI

COI

COI

COI

COI

COI

COI

COI

COI

COI

COI

COI

COI

COI

COI

## Data Availability

All numerical data backing the results are summarized in the online data supplement accompanying this preprint. Raw databases are available upon request after publication in a peer-reviewed journal.

## Author’s contributions

R.F.I. performed animal experiments, generated rodent pulmonary function and quantitative morphology data, performed data analysis and drafted the manuscript. O.A.C. and L.N.N. performed animal experiments and generated rodent pulmonary function data. B.L.C. performed animal experiments, specimen processing and immunostaining. N.S.B. coordinated multicentric biopsy collection and processing, and generated quantitative morphology data. F.J.G.B., T.B.G., C.M.M., A.L.V., A.T., V.P., C.M.R., Q.A.H., J.G.M. and P.B. recruited subjects, provided bronchial biopsies and collected clinical data. D.R.B. conceived and supervised the work and generated the final manuscript. All coauthors critically reviewed the manuscript.

## References

1. Wiggs BR, Moreno R, Hogg JC, Hilliam C, Pare PD. A model of the mechanics of airway narrowing. J Appl Physiol 1990;69:849–860.

2. Sapienza S, Du T, Eidelman DH, Wang NS, Martin JG. Structural changes in the airways of sensitized brown Norway rats after antigen challenge. Am Rev Respir Dis 1991;144:423–427.

3. Lambert RK, Wiggs BR, Kuwano K, Hogg JC, Pare PD. Functional significance of increased airway smooth muscle in asthma and COPD. J Appl Physiol 1993;74:2771–2781.

4. Salmon M, Walsh DA, Huang T-J, Barnes PJ, Leonard TB, Hay DWP, Chung KF. Involvement of cysteinyl leukotrienes in airway smooth muscle cell DNA synthesis after repeated allergen exposure in sensitized Brown Norway rats. Br J Pharmacol 1999;127:1151–1158.

5. Bousquet J, Jeffery PK, Busse WW, Johnson M, Vignola AM. Asthma. From bronchoconstriction to airways inflammation and remodeling. Am J Respir Crit Care Med 2000;161:1720–1745.

6. Leigh R, Ellis R, Wattie J, Southam DS, de Hoogh M, Gauldie J, O’Byrne PM, Inman MD. Dysfunction and Remodeling of the Mouse Airway Persist after Resolution of Acute Allergen-Induced Airway Inflammation. Am J Respir Cell Mol Biol 2002;27:526–535.

7. Henderson WRJ, Tang L, Chu S, Tsao S, Chiang GKS, Jones F, Jonas M, Pae C, Wang H, Chi EY. A Role for Cysteinyl Leukotrienes in Airway Remodeling in a Mouse Asthma Model. Am J Respir Crit Care Med 2002;165:108–116.

8. Shinagawa K, Kojima M. Mouse model of airway remodeling: strain differences. Am J Respir Crit Care Med 2003;168:959–967.

9. Woodruff PG, Dolganov GM, Ferrando RE, Donnelly S, Hays SR, Solberg OD, Carter R, Wong HH, Cadbury PS, Fahy JV. Hyperplasia of smooth muscle in mild to moderate asthma without changes in cell size or gene expression. Am J Respir Crit Care Med 2004;169:1001–1006.

10. Cox G, Thomson NC, Rubin AS, Niven RM, Corris PA, Siersted HC, Olivenstein R, Pavord ID, McCormack D, Chaudhuri R, Miller JD, Laviolette M. Asthma control during the year after bronchial thermoplasty. N Engl J Med 2007;356:1327–1337.

11. Ramos-Barbon D, Fraga-Iriso R, Brienza NS, Montero-Martinez C, Verea-Hernando H, Olivenstein R, Lemiere C, Ernst P, Hamid QA, Martin JG. T Cells localize with proliferating smooth muscle alpha-actin+ cell compartments in asthma. Am J Respir Crit Care Med 2010;182:317–324.

12. Ebina M, Takahashi T, Chiba T, Motomiya M. Cellular hypertrophy and hyperplasia of airway smooth muscles underlying bronchial asthma. A 3-D morphometric study. Am Rev Respir Dis 1993;148:720–726.

13. Gizycki MJ, Adelroth E, Rogers AV, O’Byrne PM, Jeffery PK. Myofibroblast involvement in the allergen-induced late response in mild atopic asthma. Am J Respir Cell Mol Biol 1997;16:664–673.

14. Salmon M, Walsh DA, Koto H, Barnes PJ, Chung KF. Repeated allergen exposure of sensitized Brown-Norway rats induces airway cell DNA synthesis and remodelling. Eur Respir J 1999;14:633–641.

15. Johnson PR, Roth M, Tamm M, Hughes M, Ge Q, King G, Burgess JK, Black JL. Airway smooth muscle cell proliferation is increased in asthma. Am J Respir Crit Care Med 2001;164:474–477.

16. Schmidt M, Sun G, Stacey MA, Mori L, Mattoli S. Identification of circulating fibrocytes as precursors of bronchial myofibroblasts in asthma. J Immunol 2003;171:380–389.

17. Hirst SJ, Martin JG, Bonacci JV, Chan V, Fixman ED, Hamid QA, Herszberg B, Lavoie JP, McVicker CG, Moir LM, Nguyen TT, Peng Q, Ramos-Barbon D, Stewart AG. Proliferative aspects of airway smooth muscle. J Allergy Clin Immunol 2004;114:S2–17.

18. Ramos-Barbon D, Presley JF, Hamid QA, Fixman ED, Martin JG. Antigen-specific CD4+ T cells drive airway smooth muscle remodeling in experimental asthma. J Clin Invest 2005;115:1580–1589.

19. Boxall C, Holgate ST, Davies DE. The contribution of transforming growth factor-beta and epidermal growth factor signalling to airway remodelling in chronic asthma. Eur Respir J 2006;27:208–229.

20. Saunders R, Siddiqui S, Kaur D, Doe C, Sutcliffe A, Hollins F, Bradding P, Wardlaw A, Brightling CE. Fibrocyte localization to the airway smooth muscle is a feature of asthma. J Allergy Clin Immunol 2009;123:376–384.

21. Pellettieri J, Sanchez Alvarado A. Cell turnover and adult tissue homeostasis: from humans to planarians. Annu Rev Genet 2007;41:83–105.

22. Arandjelovic S, Ravichandran KS. Phagocytosis of apoptotic cells in homeostasis. Nat Immunol 2015;16:907–917.

23. Herszberg B, Ramos-Barbon D, Tamaoka M, Martin JG, Lavoie JP. Heaves, an asthma-like equine disease, involves airway smooth muscle remodeling. J Allergy Clin Immunol 2006;118:382–388.

24. Fraga-Iriso R, Núñez-Naveira L, Mariñas-Pardo LA, Brienza NS, Verea H, Martin JG, Ramos-Barbón D. Role of apoptosis in airway smooth muscle remodeling in experimental asthma. Eur Respir J 2007;30:369s.

25. Fraga-Iriso R, Núñez-Naveira L, Mariñas-Pardo LA, Brienza NS, Verea H, Martin JG, Ramos-Barbón D. Airway Smooth Muscle (ASM) Cell Apoptosis is Upregulated during Remodeling in Experimental Asthma. Am J Respir Crit Care Med 2008;177:A324.

26. Fraga-Iriso R, Núñez-Naveira L, Mariñas-Pardo LA, Brienza NS, Verea H, Martin JG, Ramos-Barbón D. Papel de la apoptosis en la regulación del crecimiento del músculo liso durante la remodelación de vías respiratorias en asma experimental. Arch Bronconeumol 2008;44:11.

27. Fraga-Iriso R, Núñez-Naveira L, Mariñas-Pardo LA, Brienza NS, Verea H, Martin JG, Ramos-Barbón D. Upregulation of airway smooth muscle (ASM) cell apoptosis may limit ASM growth during airway remodeling in experimental asthma. Eur Respir J 2008;32:288s.

28. The Council of the European Communities. Council Directive of 24 November 1986 on the approximation of laws, regulations and administrative provisions of the Member States regarding the protection of animals used for experimental and other scientific purposes (86/609/EEC). Available at: http://ec.europa.eu/food/fs/aw/aw_legislation/scientific/86-609-eec_en.pdf.

29. Ministerio de la Presidencia. Real Decreto 1201/2005, de 10 de octubre, sobre protección de los animales utilizados para experimentación y otros fines científicos. Boletín Oficial del Estado núm. 252 de 21 de octubre de 2005, pág. 34367. Available at: http://www.boe.es/boe/dias/2005/10/21/pdfs/A34367-34391.pdf.

30. Kilkenny C, Browne WJ, Cuthill IC, Emerson M, Altman DG. Improving bioscience research reporting: the ARRIVE guidelines for reporting animal research. PLoS Biol 2010;8:e1000412.

31. National Institutes of Health National HeartLung and Blood Institute and National Institute of Allergy and Infectious Diseases, American Academy of Allergy and Immunology, American College of Chest Physicians and American Thoracic Society. Workshop summary and guidelines: investigative use of bronchoscopy, lavage, and bronchial biopsies in asthma and other airway diseases. J Allergy Clin Immunol 1991;88:808–814.

32. Investigative use of bronchoscopy, lavage and bronchial biopsies in asthma and other airways diseases. Eur Respir J 1992;5:115–121.

33. Jefatura del Estado. Ley 14/2007, de 3 de julio, de Investigación biomédica. Boletín Oficial del Estado núm. 159 de 4 de julio de 2007, pág. 28826. Available at: http://www.boe.es/boe/dias/2007/07/04/pdfs/A28826-28848.pdf.

34. Riffenburgh RH. Statistics in Medicine. San diego, California, US: Elsevier; 2012.

35. Marinas-Pardo L, Mirones I, Amor-Carro O, Fraga-Iriso R, Lema-Costa B, Cubillo I, Rodriguez Milla MA, Garcia-Castro J, Ramos-Barbon D. Mesenchymal stem cells regulate airway contractile tissue remodeling in murine experimental asthma. Allergy 2014;69:730–740.

36. Caserta TM, Smith AN, Gultice AD, Reedy MA, Brown TL. Q-VD-OPh, a broad spectrum caspase inhibitor with potent antiapoptotic properties. Apoptosis 2003;8:345–352.

37. Keoni CL, Brown TL. Inhibition of Apoptosis and Efficacy of Pan Caspase Inhibitor, Q-VD-OPh, in Models of Human Disease. J Cell Death 2015;8:1–7.

38. Hamann KJ, Vieira JE, Halayko AJ, Dorscheid D, White SR, Forsythe SM, Camoretti-Mercado B, Rabe KF, Solway J. Fas cross-linking induces apoptosis in human airway smooth muscle cells. Am J Physiol Lung Cell Mol Physiol 2000;278:L618–624.

39. Freyer AM, Johnson SR, Hall IP. Effects of growth factors and extracellular matrix on survival of human airway smooth muscle cells. Am J Respir Cell Mol Biol 2001;25:569–576.

40. Oltmanns U, Sukkar MB, Xie S, John M, Chung KF. Induction of human airway smooth muscle apoptosis by neutrophils and neutrophil elastase. Am J Respir Cell Mol Biol 2005;32:334–341.

41. Trian T, Benard G, Begueret H, Rossignol R, Girodet PO, Ghosh D, Ousova O, Vernejoux JM, Marthan R, Tunon-de-Lara JM, Berger P. Bronchial smooth muscle remodeling involves calcium-dependent enhanced mitochondrial biogenesis in asthma. J Exp Med 2007;204:3173–3181.

42. Solarewicz-Madejek K, Basinski TM, Crameri R, Akdis M, Akkaya A, Blaser K, Rabe KF, Akdis CA, Jutel M. T cells and eosinophils in bronchial smooth muscle cell death in asthma. Clin Exp Allergy 2009;39:845–855.

43. Gavrieli Y, Sherman Y, Ben-Sasson SA. Identification of programmed cell death in situ via specific labeling of nuclear DNA fragmentation. J Cell Biol 1992;119:493–501.

44. Lavrik IN, Golks A, Krammer PH. Caspases: pharmacological manipulation of cell death. J Clin Invest 2005;115:2665–2672.

45. Jenkins HA, Cool C, Szefler SJ, Covar R, Brugman S, Gelfand EW, Spahn JD. Histopathology of severe childhood asthma: a case series. Chest 2003;124:32–41.

46. Bossley CJ, Fleming L, Gupta A, Regamey N, Frith J, Oates T, Tsartsali L, Lloyd CM, Bush A, Saglani S. Pediatric severe asthma is characterized by eosinophilia and remodeling without T(H)2 cytokines. J Allergy Clin Immunol 2012;129:974–982 e913.

47. O’Reilly R, Ullmann N, Irving S, Bossley CJ, Sonnappa S, Zhu J, Oates T, Banya W, Jeffery PK, Bush A, Saglani S. Increased airway smooth muscle in preschool wheezers who have asthma at school age. J Allergy Clin Immunol 2013;131:1024-1032, 1032 e1021-1016.

48. Benayoun L, Druilhe A, Dombret MC, Aubier M, Pretolani M. Airway structural alterations selectively associated with severe asthma. Am J Respir Crit Care Med 2003;167:1360–1368.

49. James AL, Bai TR, Mauad T, Abramson MJ, Dolhnikoff M, McKay KO, Maxwell PS, Elliot JG, Green FH. Airway smooth muscle thickness in asthma is related to severity but not duration of asthma. Eur Respir J 2009;34:1040–1045.

50. Colucci WS. Apoptosis in the heart. N Engl J Med 1996;335:1224–1226.

