## Supplementary material for "Contractile cell apoptosis regulates airway smooth muscle remodeling in asthma": Online Data Supplement

### METHODS

##### Regulatory and Ethics standards

*See* statements in the article text.

##### Animal handling, induction of experimental asthma and treatments

Inbred female BALB/c mice (Harlan Interfauna Ibérica S.L., Barcelona, Spain) were accommodated into 2-L polycarbonate cages (3-4 animals per cage) equipped with air filter hoods, and placed into a MIKROS-AS-D filtered airflow rack (all equipment from Bioscape Ebeco GmbH and Co. KG, Castrop-Rauxel, Germany). Housing settings included 15-20 air exchange cycles per hour, 20-24ºC regular temperature, 55% ± 10% relative air humidity, 12/12-hour light cycle, and *ad libitum* access to water and egg protein-free feed (Teklad Global, Harlan Interfauna Ibérica S.L.). Animal welfare and sanitary control was monitored as per reference standards (E1). No research procedures were performed for a 7-day acclimatization period after animal transportation and delivery. By 6 weeks of age, the mice were sensitized to ovalbumin (OVA) by means of two intraperitoneal (*i.p.*) injections on days "0" and 7 respectively, each containing 10 µg OVA (Sigma-Aldrich, Madrid, Spain) adsorbed on 2 mg aluminum hydroxide suspension (50 µL Imject Alum Adjuvant; Thermo Scientific Pierce Protein Research Products, Barcelona, Spain) in 100 µL, pH 7.4 phosphate-buffered saline (PBS). From day 14, repeated airway challenge was performed by intranasal (*i.n.*) instillation with OVA (6.66 mg/mL in PBS) or PBS on a Monday-Wednesday-Friday regime. *See* the Results section in the main article text for each study design. The animals were subjected to short, light anesthesia with 2% sevoflurane (Sevorane) in an unrestrained inhalation chamber (anesthetic and vaporizer equipment from Abbott Laboratories, Madrid, Spain), and 12.5 µL of OVA solution or PBS were instilled per nostril in each session. Where pertinent, caspase blocker (3S)-5-(2,6-Difluorophenoxy)-3-[[(2S)-3-methyl-1-oxo-2-[(2-quinolinylcarbonyl)amino]butyl]ami-no]-4-oxo-pentanoic acid hydrate (QVD-OPH; MP Biomedicals through Grupo Taper S.A., Alcobendas, Madrid, Spain) was administered for *in vivo* inhibition of apoptosis. For this purpose, 1 mg QVD-OPH/kg body weight in dimethylsulphoxide (DMSO; Sigma-Aldrich), versus DMSO alone, was i.p. administered prior to each i.n. OVA or PBS instillation for the last 2 weeks of the instillations protocol.

**Mechanical ventilation, pulmonary function and biological specimen processing**

Pulmonary function variables were measured by forced oscillation technique under mechanical ventilation, 72 hours after the last i.n. instillation for each study protocol. Anesthesia was induced with 2% sevoflurane (Sevorane) and then maintained with a mixed flow of 3.5-4.5 mL/min Sevorane and 3-5 L/min oxygen. Upon verification of deep anesthesia by flexor (leg withdrawal) and corneal reflex check, the animals were tracheostomized and intubated with a FTC-100 tracheal cannula (Electromedsys EMMS, Bordon, Hants, UK). Peripheral vein catheterization was performed in the tail using human epidural anesthesia catheters (Yale™ Spinal 18G x 3½; Becton Dickinson & Co., San Agustín de Guadalix, Madrid, Spain). Neuromuscular blockade was induced with a 0.15-mL intravenous (i.v.) bolus of 0.5 mg/mL rocuronium bromide (Organon, Cornellá de Llobregat, Barcelona, Spain). Additional 0.1-mL i.v. doses were administered at 15 minutes after the initiation of mechanical ventilation and as needed through the complete procedure. The animals were connected to calibrated FlexiVent 5.0 mechanical ventilation and pulmonary function equipment (SciReq, Montreal, Canada; purchased through Electromedsys EMMS). Single-lead EKG monitoring was performed from subcutaneous electrodes (Electromedsys EMMS). Maintenance mechanical ventilation was performed at 250 $min^{-1}$ respiratory rate with a 0.12-0.14 mL/kg body weight tidal volume, resulting in 1.2-1.3 L/kg minute ventilation, and a 1-2 $cmH_2O$ positive end expiratory pressure (PEEP) was applied. Pulmonary resistance ($R_L$) and elastance measurements were generated from “snapshot” perturbation maneuvers as per single-compartment model. Following the recording of several stable baseline $R_L$ and elastance values, a methacholine bronchoprovocation test was performed. For this purpose, a dilution series of methacholine (Sigma-Aldrich) in PBS was administered at 0 (PBS diluent), 1.25, 2.5, 5 and 10 mg/mL concentrations by means of an Aeroneb ultrasonic nebulizer (Electromedsys EMMS). For each dilution, 25 µL were nebulized in 15 seconds and data acquisition was done every 20 seconds for 4 minutes to capture the $R_L$ peak. Upon completion of pulmonary function data acquisition, animal euthanasia was performed by means of an anesthesia overdose along with a 0.3 mL i.v. bolus of 1 M potassium chloride (B Braun, Rubí, Barcelona, Spain), and a 0.1 mL bolus of 1% sodium heparin (Mayne Pharma, Madrid, Spain) followed. Bronchoalveolar lavage (BAL) was performed through the intratracheal cannula with 5 mL PBS, instilled and retrieved in 1-mL fractions by means of a 3-way stopcock. The cellular fraction was separated by centrifugation at 458·g. Total viable leukocytes were counted by trypan blue dye exclusion in a Neubauer hemacytometer chamber from the cell pellet resuspended in 1 mL PBS. The cell concentration was then adjusted to 400,000 cells/mL, and the cell suspension was cytocentrifuged at 28·g for 6 minutes (Universal 32 cytocentrifuge; Hettich, Tuttlingen, Germany) on adherent microscope slides (SuperFrost™ Plus, Sigma-Aldrich) so as to deposit 60,000 cells per the slide. The slides were air dried for 20 minutes, fixed in methanol (VWR International Eurolab, Llinars del Vallès, Barcelona, Spain) for 10 minutes, air dried again, and stained with Wright-Giemsa (VWR International Eurolab) for leukocyte differential counts. Following BAL, a mid thoraco-abdominal incision was done, the inferior vena cava and abdominal aorta were sectioned, and the pulmonary vascular circuit was flushed with PBS through injection into the right ventricle. The cardiopulmonary block was then dissected out and connected through the tracheal cannula to a recirculating pump driving "10% formalin" (4% formaldehyde in PBS; stock formaldehyde from Panreac, Castellar del Vallès, Barcelona, Spain) for 24-hour fixation at 20 $cmH_2O$ standard pressure. The lungs were then sagittally sliced, immersed in 70% ethanol (Panreac) into histological casettes, and run overnight through tissue processor equipment (Leica Microsystems, Barcelona, Spain) for paraffin embedding. Four-µm thick tissue sections were cut and placed on SuperFrost™ Plus slides for further processing.

**Bronchial biopsy subjects**

Subjects with persistent asthma where classified as severe as per the American Thoracic Society Workshop on Refractory Asthma (E2), or as moderate as per criteria reported elsewhere (E3). Subjects with asthma classed as intermittent were managed in therapeutic Step 1 (on-demand short-acting $\beta_2$-agonist bronchodilator) as per the Global INitiative for Asthma (GINA) guidelines (E4). For severe asthmatics, at least one of the two following major criteria had to be met: (a) treatment with daily oral corticosteroids for more than 50% of the previous 12 months or (b) treatment with high-dose inhaled steroid (≥1000 mg of fluticasone or equivalent per day) and at least one other add-on therapy (long-acting $\beta_2$-agonist, leukotriene receptor antagonist, or theophylline) continuously over the previous 12 months. In addition, at least 2 or more minor criteria had to be present: (a) daily use of short-acting $\beta_2$-agonist; (b) persistent airflow obstruction, as per prebronchodilator $FEV_1$ of less than 70% and an $FEV_1/FVC$ ratio of less than 80% of predicted value; (c) 1 or more urgent care visits in the previous 12 months; (d) 3 or more corticosteroid bursts in the previous 12 months; (e) prompt deterioration with less than 25% dose reduction of steroids; or (f) a near-fatal asthma event in the last 3 years. For the moderate asthmatics, all of the following criteria had to be met: a) well-controlled asthma on at least 200 mg/day of fluticasone propionate or equivalent but not exceeding 1000 mg/day, with or without the use of a long-acting $\beta_2$-agonist; (b) 2 or less corticosteroid bursts in the previous 12 months and none in the past 3 months (the total number of days taking oral corticosteroids could not exceed 30 days in the previous 12 months); (c) $FEV_1$ >70% of predicted value and >90% of the individual's best from the previous 2 years; and (d) no more than one nonscheduled urgent care or clinic visit in the previous 12 months. As for the control group, these were subjects without asthma or other conditions involving airway inflammation, with a basal $FEV_1 \geq 80\%$ of its predicted value, no medical therapy that could affect airway physiology, and normal airways at bronchoscopic examination. Bronchial biopsies were collected from areas with no visible pathology upon bronchoscopic examination, and distant from any area of focal pathology under study. A bronchoscopy was performed under indications such as laser treatment of tracheal stenosis, ruling out malignancy on a prior tuberculous scar, or the investigation of conditions including a focal lung infiltrate with anterior mediastinal lymphadenopathy (suspected tuberculosis), mediastinal lymphadenopathies (suspected sarcoidosis), erythema nodosum with mediastinal lymphadenopathy (suspected sarcoidosis), hemoptysis of unknown origin, and the study of chronic cough with asthma and gastroesophageal reflux ruled out.

Exclusion criteria were: (i) any disability preventing from understanding and signing the informed consent form; (ii) any general contraindication for bronchoscopy and/or bronchial biopsy as per standard clinical practice; and (iii) the presence of any known pulmonary disease (except for the focal condition under investigation where applicable), or any comorbidity that might affect asthma disease activity or airway physiology, immunobiology and/or histopathology overall, such as HIV infection, metastatic cancer, congestive heart failure or other conditions upon investigator's judgment. All subjects were nonsmokers or ex-smokers of at least 2 years with a smoking history of less than 5 packs-years.

**Bronchial biopsy collection and processing**

Bronchial biopsies for biobank collection and investigational purposes were obtained in a multicenter fashion following published guidelines and recommendations (*see* Methods section in the article text). Collaborating sites within the Spanish network were issued kits providing all necessary documents and expendables for biopsy collection, packing and shipping. The centers in Canada and France directly provided unstained tissue sections on microscope slides, from tissue blocks kept at their own site collections. Biopsy collection procedures were uniform across all centers. Each kit issued to centers in Spain contained: an autoclavable, reusable Olympus 35C biopsy forceps; a set of tight-seal screw-cap cryotubes (Fisher Scientific, Barcelona, Spain) preloaded with 4% formaldehyde ("10% formalin"); approved shipping materials for biological specimens in compliance with the International Air Transport Association (IATA) regulations, pre-labeled with the destination address; a document filing binder; informed consent forms; case report form ("CRF") documents; a DHL courier account number with pickup contact; and cell phone access numbers to the *Bronchial Biopsy Biobank* project manager and principal investigator for shipping alert. Biopsies were collected from the carinae of the first subsegmental bronchial generation, unilaterally, and from the opposite lung or lung regions distant from any focal pathology under diagnostic investigation where applicable. Upon extraction, biopsies were immediately released into fixative-containing cryotubes, the tubes labeled with the subject's anonymization code and a sequential biopsy number, and shipped. The biopsies were fixed during overnight shipment at room temperature. Upon receipt, at 24 hours after the recorded biopsy collection time, tissue fixation was stopped by transferring the biopsies to 70% ethanol and the specimens underwent tissue processing for paraffin embedding through the following night. Five-µm thick tissue sections were cut and laid on SuperFrost Plus adherent slides for staining and analysis.

***In situ* specific detections and quantitative morphology**

Prior to any procedure, tissue sections were deparaffined and rehydrated through xylene, ethanol and Tris-buffered saline (TBS; compounds from Sigma-Aldrich). Apoptosis was detected by means of an ApopTag® Peroxidase In Situ Apoptosis Detection Kit (Chemicon® International/Millipore, Madrid, Spain), as per manufacturer's instructions. For this purpose, the tissues were pretreated with 20 µg/mL proteinase K (Sera Laboratories International Ltd., West Sussex, UK) for 15 minutes at 37°C inside a humid chamber, and endogenous peroxidase activity was quenched with commercial peroxidase blocking solution (Dako, Sant Just Desvern, Barcelona, Spain). Signal development was done with diaminobenzidine (DAB)-nickel (Vector Laboratories through Palex Medical, Sant Cugat del Vallès, Barcelona, Spain). Active caspase-3 was detected using rabbit monoclonal IgG (Millipore; product ID 04-439) primary antibody at 1:50 dilution in TBS with 2.5% goat serum (Vector Laboratories) and 0.2% Tween-20 (Sigma-Aldrich). An antigen retrieval pretreatment was done with

1 mg/mL trypsin (Sigma-Aldrich) at 37°C for 30 minutes, followed by membrane permeabilization with 0.2% Triton X-100 (Sigma-Aldrich) in TBS for 20 minutes, blocking with 5% goat serum, and endogenous avidin-biotin blockade with Avidin/Biotin Blocking Kit (Vector Laboratories). The detection system consisted of: (i) a biotinylated goat anti-rabbit secondary antibody (Vector Laboratories) at 5 µg/mL concentration with 5% mouse serum (Sigma-Aldrich) and 02% Tween-20 in TBS; (ii) an avidin-biotin alkaline phosphatase complex (ABC-AP, Vector Laboratories); and (iii) signal development with 5-bromo-4-chloro-3-indolyl phosphate/nitroblue tetrazolium (BCIP/NBT) chromogen (Vector Laboratories). Cleaved poly (ADP-ribose) polymerase (c-PARP) was detected with rabbit monoclonal (clone E51) primary antibody (Abcam, Cambridge, UK; product ID ab32064) at 1:50 dilution in TBS with 2.5% goat serum and 0.2% Tween-20. The specimens were pretreated through a heat antigen retrieval procedure in pH 6 citrate buffer (Dako) using Antigen Retriever 2100 equipment (Antigen-Retriever, Southhampton, UK), followed by membrane permeabilization, blocking with 5% goat serum, and endogenous avidin-biotin blockade. The detection system was as for active caspase-3 on mouse tissue sections. In the case of human biopsy sections, the secondary antibody diluent contained 5% human serum (Sigma-Aldrich) instead of mouse serum. After development of TUNEL, active caspase-3 or c-PARP detection, the signals were colocalized with smooth muscle α-actin (α-SMA), using mouse monoclonal primary antibody clone 1A4 (Sigma-Aldrich) at 2 µg/mL working concentration. In the case of mouse tissue sections, a "mouse-on-mouse" tissue pretreatment and detection system was employed (M.O.M. Kit, Vector Laboratories) as per manufacturer's instructions, along with ABC-AP complex. On human bronchial biopsy sections, the tissue was blocked with 5% horse serum (Vector Laboratories) in TBS with 0.2% Tween-20, the primary antibody was applied in 2.5% horse serum with 0.2% Tween-20 in TBS, and this was followed by biotinylated horse anti-mouse secondary antibody (Vector Laboratories) at 5 µg/mL concentration in TBS with 5% human serum and 0.2% Tween-20, and ABC-AP complex. For both mouse and human coimmunostaining, the α-SMA signal was developed with Vector-Red Alkaline Phosphatase Substrate Kit (Vector Laboratories). All colocalization procedures were followed by counterstaining with 0.05% methyl green in 0.1 M, pH 4 sodium acetate (Sigma-Aldrich). The specimens were then dehydrated through 1-butanol and xylene, and mounted with VectaMount permanent medium (Vector Laboratories). In the case of α-SMA detection for quantitation of AwCT in the murine models, the primary 1A4

antibody was detected with an f(ab)` anti-mouse IgG2a labeled with Alexa-488 (Zenon® Alexa Fluor® 488 Mouse IgG2a Labeling Kit, Molecular Probes/Invitrogen, Madrid, Spain). The specimens were counterstained with 4',6-diamidino-2-phenylindole (DAPI) using SelectFX™ Nuclear Labeling Kit (Molecular Probes/Invitrogen), then fixed in 4% paraformaldehyde (Sigma-Aldrich) and permanently mounted in ProLong® Gold mounting media (Molecular Probes/Invitrogen).

Quantitative morphology was blindly performed on coded preparations to calculate murine AwCT mass, human airway smooth muscle mass, and the frequency of TUNEL$^+$α-SMA$^+$, active caspase-3$^+$α-SMA$^+$ and c-PARP$^+$α-SMA$^+$ cells in mouse airways and human bronchial biopsy sections. Stereological measurements on the basis of volumetric probes employing thick tissue sections (E5) were technically unfeasible due to the tissue pretreatments needed for specimen processing (particularly proteinase K and trypsin enzymatic treatments). Such treatments, when applied to rehydrated, 25 μm-thick sections as a minimum thickness recommended for stereology, led to tissue detachment or fracture, and irregular thickness of the processed preparations, thus rendering reliable stereology unfeasible. Quantitative morphology was therefore performed as per alternative established methods (E6-E8). AnalySISD® 5.0 (Soft Imaging System GmbH, Münster, Germany) or Image-J software (National Institutes of Health, Bethesda, MD, U.S.) was employed for quantitative image analysis. Calibrated microscopy equipment with a digital camera (Olympus, L'Hospitalet de Llobregat, Barcelona, Spain) was employed for image acquisition. In the murine lung sections, all cross-sectioned airways (with the long diameter not exceeding 1.5-fold the short diameter) were systematically sampled and, in the human bronchial specimens, the complete biopsy section was captured. Whether from murine or human specimens, the α-SMA signal was digitally extracted, converted to grayscale and delimited with regions of interest ("R.O.I.") to exclude vascular smooth muscle and epithelial noise where pertinent, and the surface area of the resulting α-SMA signal was then calculated. Cell counts were performed at high resolution under direct microscope view (using a 40x objective or a 100x objective with oil immersion as needed). All measurements were referenced for normalization to the basement membrane perimeter squared in the case of the mouse lung sections, or to the section surface area for the bronchial biopsies.

**ARRIVE reporting items compliance**

This section provides supplemental information to complete ARRIVE Guidelines reporting items. For each set of experiments, the study design is reported in full detail the corresponding Results section and illustrated in Figures 1 and 2. The experimental unit conforming the data distributions is each individual animal. The size of each experimental group is reported in the supplemental data tables along with the numerical data pertaining to the distribution statistics and size of effect estimates. *A priori* sample size calculation was not possible due to the unknown variability of the outcome distributions and expected size of effects. Group size was set within the range of usual experimental asthma reports in the literature as n=8 by default and n=11 in the OVA+DMSO and OVA+QVD-OPH groups due to their large $R_L$ variability. The assignment of animals to the experimental groups was randomized by sequential cage sampling. All animals in the same cage received the same treatment to avoid marking of individual animals and prevent error. The animals were included in the experiments sequentially by cage, alternating cages by treatment, with no preset exclusion criteria. No potential confounders were identified attributable to animal supply, handling, study design, treatments, or sequence of procedures.

### RESULTS (Supplemental Data)

**Table E1. Airway hyperresponsiveness at early-disease cutoff**

| | **MCh mg/mL** | **$R_L$ $cmH_2O \cdot s/mL$** | ***P*** | | **Mean Δ [ 95% CI] $cmH_2O \cdot s/mL$** | |
|---|---|---|---|---|---|---|
| **CTRL** (n = 8) | **Baseline** | 1.02 ± 0.07 | | - | | - |
| | **PBS** | 1.27 ± 0.23 | - | - | - | - |
| | **1.25** | 1.16 ± 0.09 | - | - | - | - |
| | **2.5** | 1.43 ± 0.14 | - | - | - | - |
| | **5** | 1.73 ± 0.22 | - | - | - | - |
| | **10** | 2.56 ± 0.54 | - | - | - | - |
| **OVA** (n = 8) | **Baseline** | 0.82 ± 0.05 | - | - | - | - |
| | **PBS** | 0.77 ± 0.05 | - | - | - | - |
| | **1.25** | 1.01± 0.08 | - | - | - | - |
| | **2.5** | 1.53 ± 0.18 | - | - | - | - |
| | **5** | 3.70 ± 0.84 | 0.047[a] | <0.001[b] | 2.03 [0.46; 4.00 ][a] | 2.88 [1.53][b] |
| | **10** | 5.70 ± 0.56 | 0.002[a] | <0.001[b] | 3.13 [1.43; 4.84 ][a] | 5.16 [3.78][b] |

$R_L$ values are mean ± SEM for each MCh concentration. *P* values are quoted where significant, and the size of the corresponding effect is estimated by the 95% CI of the mean increment (**Δ**) or its lower bound in the case of one-tailed contrast versus intra-group baseline $R_L$. [a]: OVA versus CTRL. [b]: versus intra-group baseline $R_L$.

**Table E2. AwCT mass, early and long-term disease cutoffs**

| | | AwCT mass | *P* | | Mean Δ [ 95% CI] | |
|---|---|---|---|---|---|---|
| Early cutoff | CTRL (n = 8) | 1.04 ± 0.28 | | | | |
| | OVA (n = 8) | 4.47 ± 0.38 | <0.001[a] | 0.016[b] | 3.43 [2.14, 4.71][a] | 1.85 [1.07, 2.63][b] |
| Long-term cutoff | CTRL (n = 8) | 1.02 ± 0.07 | | | | |
| | OVA (n = 8) | 2.62 ± 0.18 | <0.001[a] | | 1.59 [1.17, 2.02][a] | |

AwCT (dimensionless) is expressed as mean ± SEM. *P* values are quoted where significant. The size of effects is estimated by the 95% CI of the mean increment (**Δ**). [a]: OVA versus CTRL. [b]: OVA groups, early versus long-term disease cutoffs.

**Table E3. AwCT cell apoptosis at early and long-term disease cutoffs**

| | | Mean ± SEM cells/mm$^2$ | *P* | Mean Δ [95% CI] cells/mm$^2$ | |
|---|---|---|---|---|---|
| TUNEL$^+$ | CTRL-Early | 0.79 ± 0.13 | | | |
| | OVA-Early | 0.50 ± 0.13 | - | - | |
| | CTRL-Long | 0.60 ± 0.16 | | | |
| | OVA-Long | 3.59 ± 0.57 | <0.001[a,b] | 2.99 [2.04; 3.93][a] | 3.09 [2.17; 4.00][b] |
| Casp-3$^+$ | CTRL-Early | 0.32 ± 0.11 | | | |
| | OVA-Early | 0.20 ± 0.05 | - | - | |
| | CTRL-Long | 0.32 ± 0.10 | | | |
| | OVA-Long | 1.25 ± 0.36 | 0.006[a,b] | 0.93 [0.32; 1.54][a] | 1.05 [0.44; 1.66][b] |

Values are the frequency of α-SMA$^+$TUNEL$^+$ and α-SMA$^+$active caspase-3 (casp-3)$^+$ cells, corrected by airway size as per basement membrane perimeter squared ($P_{BM}^2$). *P* values are quoted where significant. The 95% CIs of the mean increment (Δ) estimate the size of the effects. [a]: OVA versus CTRL group; [b]: OVA groups, early versus long-term cutoffs. Group sizes as in Table E2.

**Table E4. $R_L$, QVD-OPH apoptosis inhibition model**

| | MCh (mg/mL) | $R_L$, mean ± SEM [Range] $cmH_2O·s/mL$ | *P* | Mean Δ [95% CI] $cmH_2O·s/mL$ |
|---|---|---|---|---|
| PBS + DMSO (n = 8) | Baseline | 0.59 ± 0.01 [0.54 - 0.65] | | |
| | PBS | 0.55 ± 0.02 [0.49 - 0.64] | 0.877[d] | - |
| | 1.25 | 0.90 ± 0.13 [0.58 - 1.65] | 0.370[d] | - |
| | 2.5 | 1.39 ± 0.21 [0.78 - 2.44] | 0.014[d] | 0.80 [0.16][d] |
| | 5 | 2.27 ± 0.35 [0.91 - 3.70] | <0.001[d] | 1.68 [1.03][d] |
| | 10 | 2.70 ± 0.23 [1.67 - 3.67] | <0.001[d] | 2.11 [1.47][d] |
| PBS + QVD OPH (n = 8) | Baseline | 0.67 ± 0.04 [0.54 - 0.82] | 0.264[a] | - |
| | PBS | 0.64 ± 0.04 [0.48 - 0.82] | 0.245[a]; 0.869[d] | - |
| | 1.25 | 0.88 ± 0.04 [0.75 - 1.15] | 0.994[a]: 0.497[d] | - |
| | 2.5 | 1.27 ± 0.13 [0.86 - 1.85] | 0.958[a]; 0.047[d] | 0.60 [0.007][d] |
| | 5 | 1.59 ± 0.28 [0.93 - 3.15] | 0.450[a]; 0.002[d] | 0.92 [0.32][d] |
| | 10 | 2.33 ± 0.32 [1.43 - 3.61] | 0.777[a]; <0.001[d] | 1.66 [1.06][d] |
| OVA + DMSO (n = 11) | Baseline | 1.26 ± 0.48 [0.56 - 6.01] | 0.530[a]; 0.624[b] | - |
| | PBS | 1.38 ± 0.59 [0.53 - 7.16] | 0.245[a]; 0.604[b]; 0.794[d] | - |
| | 1.25 | 1.60 ± 0.60 [0.68 - 7.53] | 0.678[a]; 0.648[b]; 0.708[d] | - |
| | 2.5 | 2.25 ± 0.62 [0.91 - 7.86] | 0.581[a]; 0.451[b]; 0.402[d] | - |
| | 5 | 3.39 ± 0.74 [1.49 - 8.53] | 0.485[a]; 0.116[b]; 0.047[d] | 2.21 [0.02][d] |
| | 10 | 6.05 ± 0.94 [2.65 - 12.38] | 0.023[a]; 0.012[b]; <0.001[d] | 3.35 [0.45, 6.24][a] 3.72 [0.79, 6.65][b] 4.79 [2.61][d] |
| OVA + QVD OPH (n = 11) | Baseline | 4.88 ± 4.10 [0.68 - 45.83] | 0.222[a]; 0.231[b]; 0.261[c] | - |
| | PBS | 5.79 ± 5.10 [0.54 - 56.81] | 0.230[a]; 0.238[b]; 0.270[c]; 0.788[d] | - |
| | 1.25 | 4.22 ± 2.88 [0.57 - 32.62] | 0.186[a]; 0.184[b]; 0.253[c]; 0.867[d] | - |
| | 2.5 | 5.04 ± 3.35 [0.65 - 38.29] | 0.210[a]; 0.195[b]; 0.294[c]; 0.829[d] | - |
| | 5 | 13.26 ± 6.02 [0.86 - 50.96] | 0.037[a]; 0.028[b]; 0.041[c]; 0.252[d] | 10.99 [0.67, 21.30][a] 11.67 [1.36, 21.98][b] 9.87 [0.40, 19.33][c] |
| | 10 (n = 9)* | 7.74 ± 2.67 [1.47 - 26.98] | 0.025[a]; 0.016[b]; 0.399[c]; 0.670[d] | 5.03 [0.69, 9.38][a] 5.41 [1.06, 9.76][b] |

$R_L$ data are mean ± SEM. The 95% CIs of the mean increment (Δ) estimate the size of the effects where significant. Mch: methacholine. [a]: versus [i.n. PBS + i.p. DMSO]; [b]: versus [i.n. + i.p. QVD-OPH]; [c]: versus [i.n. OVA + i.p. DMSO]; [d]: versus intra-group baseline $R_L$, one-tailed ([lower-bound CI value]). Group size applies to all measurements except otherwise specified. *: Two animals died at this point from airway closure, unresponsive to suction and ventilator-managed maneuvering. These animals were processed for biological specimen analyses and not excluded from the data sets in Tables E5 to E7 and Figure E1.

**Table E5. AwCT mass, QVD-OPH apoptosis inhibition model**

| | Mean ± SEM | *P* | Mean Δ [95% CI] |
|---|---|---|---|
| **i.n. PBS + i.p. DMSO** (n = 8) | 0.85 ± 0.11 | | |
| **i.n. PBS + i.p. QVD-OPH** (n = 8) | 1.43 ± 0.15 | 0.003[a] | 0.58 [0.24, 0.93][a] |
| **i.n. OVA + i.p. DMSO** (n = 11) | 2.34 ± 0.13 | <0.001[a]<br><0.001[b] | 1.49 [1.13, 1.84][a]<br>0.91 [0.56, 1.25][b] |
| **i.n. OVA + i.p. QVD-OPH** (n = 11) | 3.80 ± 0.31 | <0.001[a]<br><0.001[b]<br><0.001[c] | 2.95 [2.57, 3.33][a]<br>2.37 [2.00, 2.74][b]<br>1.46 [1.08, 1.84][c] |

AwCT mass is dimensionless. The 95% CIs of the mean increment (Δ) estimate the size of the effect where differences are significant. [a]: versus [i.n. PBS + i.p. DMSO]; [b]: versus [i.n. PBS + i.p. QVD-OPH]; [c]: versus [i.n. OVA + i.p. DMSO].

**Table E6. AwCT cell apoptosis in QVD-OPH model**

| | | Mean ± SEM cells/mm$^2$ | *P* | Mean Δ [95% CI] cells/mm$^2$ |
|---|---|---|---|---|
| TUNEL$^+$ | **i.n. PBS + i.p. DMSO** (n = 8) | 0.24 ± 0.09 | | |
| | **i.n. PBS + i.p. QVD-OPH** (n = 8) | 0.07 ± 0.07 | 0.523[a] | - |
| | **i.n. OVA + i.p. DMSO** (n = 11) | 1.31 ± 0.35 | 0.007[a]<br>0.002[b] | 1.07 [0.30, 1.83][a]<br>1.24 [0.47, 2.00][b] |
| | **i.n. OVA + i.p. QVD-OPH** (n = 11) | 0.97 ± 0.15 | 0.060[a]<br>0.022[b]<br>0.336[c] | -<br>0.90 [0.13, 1.66][b]<br>- |
| Casp-3$^+$ | **i.n. PBS + i.p. DMSO** (n = 8) | 0.70 ± 0.12 | | |
| | **i.n. PBS + i.p. QVD-OPH** (n = 8) | 0.77 ± 0.08 | 0.719[a] | - |
| | **i.n. OVA + i.p. DMSO** (n = 11) | 1.05 ± 0.15 | 0.066[a]<br>0.141[b] | -<br>- |
| | **i.n. OVA + i.p. QVD-OPH** (n = 11) | 0.55 ± 0.12 | 0.414[a]<br>0.232[b]<br>0.005[c] | -<br>-<br>-0.50 [-0.84, -0.16][c] |
| c-PARP$^+$ | **i.n. PBS + i.p. DMSO** (n = 8) | 1.81 ± 0.14 | | |
| | **i.n. PBS + i.p. QVD-OPH** (n = 8) | 1.20 ± 0.15 | 0.048[a] | -0.61 [-1.22, -0.006][a] |
| | **i.n. OVA + i.p. DMSO** (n = 11) | 2.19 ± 0.36 | 0.326[a]<br>0.013[b] | -<br>0.99 [0.22, 1.76][b] |
| | **i.n. OVA + i.p. QVD-OPH** (n = 11) | 1.57 ± 0.23 | 0.509[a]<br>0.330[b]<br>0.084[c]* | -<br>-<br>-0.62 [-1.32, -0.09][c]* |

Values express the frequency of α-SMA$^+$TUNEL$^+$, α-SMA$^+$active caspase-3 (casp-3)$^+$ and α-SMA$^+$c-PARP$^+$ cells referenced to airway basement membrane perimeter squared ($P_{BM}^2$). The 95% CIs of the mean increment (Δ) estimate the size of the effects for statistically significant differences, or borderline significant differences for relevant comparisons as indicated. [a]: versus [i.n. PBS + i.p. DMSO]; [b]: versus [i.n. PBS + i.p. QVD-OPH]; [c]: versus [i.n. OVA + i.p. DMSO]; *: borderline statistical significance.

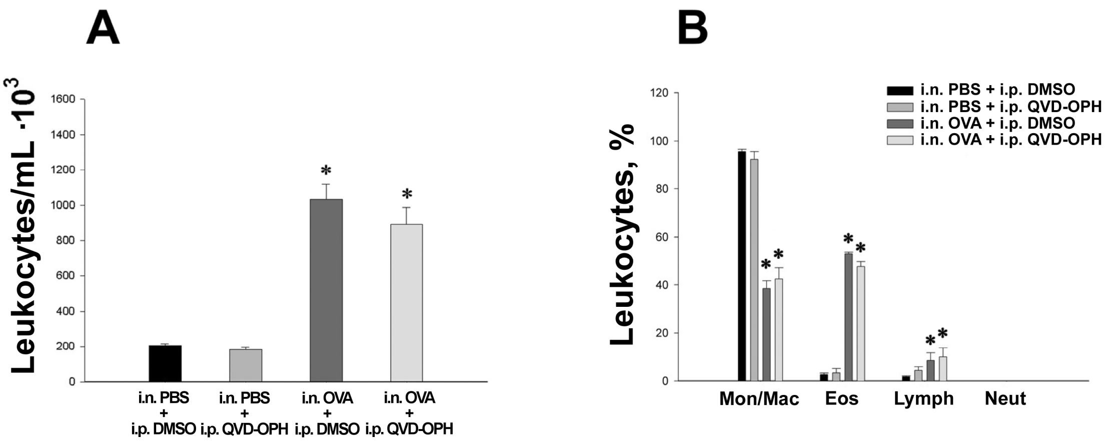


**Figure E1. Total (A) and differential (B) leukocyte counts in BAL.** Data are presented as mean and SEM. Mon/Mac: monocytes/macrophages pooled count. Eos: eosinophils. Lymph: lymphocytes. Neut: neutrophils. *: *P*<0.05 versus [i.n. PBS + i.p. DMSO] and [i.n. PBS + i.p. QVD-OPH].

**Table E7 (A). Total BAL leukocyte counts**

| | | **Mean ± SEM** Cells/mL · $10^3$ | ***P*** | **Δ [95% CI]** Cells/mL · $10^3$ |
|---|---|---|---|---|
| **Total leukocytes** | **i.n. PBS + i.p. DMSO** (n = 8) | 207.50 ± 10.51 | - | - |
| | **i.n. PBS + i.p. QVD-OPH** (n = 8) | 182.50 ± 16.29 | 0.815[a] | - |
| | **i.n. OVA + i.p. DMSO** (n = 11) | 1031.90 ± 104.23 | <0.001[a] | 824.37 [607.73, 1041.00][a] |
| | | | <0.001[b] | 849.37 [632.73, 106.70][b] |
| | **i.n. OVA + i.p. QVD-OPH** (n = 11) | 894.00 ± 106.58 | <0.001[a] | 686.50 [485.17, 887.83][a] |
| | | | <0.001[b] | 711.50 [510.16, 912.83][b] |
| | | | 0.172[c] | - |

See legend under table partition (B).

**Table E7 (B). Differential BAL leukocyte counts**

| | | **Mean ± SEM** % | ***P*** | **Δ [95% CI]** Cells/mL · $10^3$ |
|---|---|---|---|---|
| **Monocytes/ Macrophages** | **i.n. PBS + i.p. DMSO** | 95.60 ± 0.67 | - | - |
| | **i.n. PBS + i.p. QVD-OPH** | 92.33 ± 2.59 | 0.600[a] | - |
| | **i.n. OVA + i.p. DMSO** | 38.46 ± 2.76 | <0.001[a] | -57.14 [-68.61, -45,67][a] |
| | | | <0.001[b] | -53.87 [-65.34, -42.40][b] |
| | **i.n. OVA + i.p. QVD-OPH** | 42.37 ± 4.04 | <0.001[a] | -53.22 [-64.70, -41.75][a] |
| | | | <0.001[b] | -49.96 [-61.43, -38.48][b] |
| | | | 0.428[c] | - |
| **Eosinophils** | **i.n. PBS + i.p. DMSO** | 2.66 ± 0.46 | - | - |
| | **i.n. PBS + i.p. QVD-OPH** | 3.33 ± 1.26 | 0.898[a] | - |
| | **i.n. OVA + i.p. DMSO** | 53.08 ± 2.77 | <0.001[a] | 50.41 [40.79, 60.04][a] |
| | | | <0.001[b] | 49.75 [40.12, 59.38][b] |
| | **i.n. OVA + i.p. QVD-OPH** | 47.62 ± 3.24 | <0.001[a] | 44.96 [35.33, 54.59][a] |
| | | | <0.001[b] | 44.29 [34.66, 53.91][b] |
| | | | 0.194[c] | - |
| **Lymphocytes** | **i.n. PBS + i.p. DMSO** | 1.73 ± 0.25 | - | - |
| | **i.n. PBS + i.p. QVD-OPH** | 4.33 ± 1.40 | 0.286[a] | - |
| | **i.n. OVA + i.p. DMSO** | 8.46 ± 0.62 | 0.005[a] | 6.72 [2.28, 11.17][a] |
| | | | 0.249[b] | - |
| | **i.n. OVA + i.p. QVD-OPH** | 9.95 ± 1.67 | 0.001[a] | 8.22 [3.78, 12.67][a] |
| | | | 0.015[b] | 5.62 [1.18, 10.06][b] |
| | | | 0.433[c] | - |

Values for differential leukocyte counts are expressed as mean percentages and SEM. Neutrophils were only detected in the [i.n. OVA + i.p. QVD-OPH] group with a frequency of 4.04% ± 0.03%. The 95% CIs of the mean increment (Δ) estimate the size of the effect where differences are significant. [a]: versus [i.n. PBS + i.p. DMSO]; [b]: versus [i.n. PBS + i.p. QVD-OPH]; [c]: versus [i.n. OVA + i.p. DMSO].

**Table E8. Demographics and clinical data**

| | | **Age** mean ± SEM [range] | **Gender** female:male | **Atopy** Yes:No | **FEV1, % predicted** mean ± SEM | **FEV1/FVC ratio, %** mean ± SEM |
|---|---|---|---|---|---|---|
| | **Controls (n=13)** | 51 ± 2.3 [32-62] | 7:6 | 1:12 | 88.32 ± 5.51 | 80.20 ± 2.95 |
| **Asthmatics** | **Intermittent (n=7)** | 29 ± 4.0 [20-49] | 5:2 | 6:1 | 99.41 ± 3.70 | 82.66 ±1.25 |
| **Asthmatics** | **Moderate (n=33)** | 42 ± 2.7 [18-74] | 18:15 | 25:8 | 91.38 ± 2.41 | 75.84 ± 1.16 |
| **Asthmatics** | **Severe (n=19)** | 49 ± 3.4 [17-69] | 11:8 | 17:2 | 64.74 ± 4.62 | 63.23 ± 3.21 |

Age was homogeneously distributed among controls, and moderate and severe asthmatics. The subjects with intermittent asthma were significantly younger than the other groups ($P$ = 0.001, $P$ = 0.028 and $P$ = 0.002 versus controls, moderate asthmatics and severe asthmatics, respectively). Gender was homogeneously distributed. Atopy was significantly more frequent in the severe asthmatics ($P$<0.001). $FEV_1$ and the $FEV_1$/FVC ratio were preserved in the intermittent and moderate asthmatics, and were significantly decreased in the severe asthmatics ($P$<0.001 versus all other groups for both parameters).

**Table E9. Airway smooth muscle mass, and frequency of α-SMA⁺TUNEL⁺ and α-SMA⁺c-PARP⁺ cells in bronchial biopsies**

| | | Mean ± SEM | *P* | Δ [95% CI] |
|---|---|---|---|---|
| ASM mass | Controls | 0.085 ± 0.013 | | |
| | Intermittent asthma | 0.161 ± 0.024 | 0.087[a]* | 0.076 [-0.011, 0.163][a]* |
| | Moderate asthmatics | 0.179 ± 0.019 | 0.003[a] | 0.094 [0.033, 0.155][a] |
| | | | 0.644[b] | - |
| | Severe asthmatics | 0.242 ± 0.019 | <0.001[a] | 0.156 [0.089, 0.223][a] |
| | | | 0.056[b]* | 0.080 [-0.002, 0.162][b]* |
| | | | 0.023[c] | 0.062 [0.009, 0.116][c] |
| TUNEL cells/mm² | Controls | 1.273 ± 0.418 | | |
| | Intermittent asthma | 3.095 ± 0.959 | 0.152[a] | - |
| | Moderate asthmatics | 2.381 ± 0.449 | 0.961[a] | - |
| | | | 0.465[b] | - |
| | Severe asthmatics | 2.351 ± 0.505 | 0.206[a] | - |
| | | | 0.475[b] | - |
| | | | 0.965[c] | - |
| c-PARP cells/mm² | Controls | 4.456 ± 0.606 | | |
| | Moderate asthmatics | 3.618 ± 1.526 | 0.829[a] | - |
| | Severe asthmatics | 13.677 ± 4.042 | 0.008[a] | 9.221 [2.685, 15.757][a] |
| | | | 0.021[c] | 10.059 [1.651, 18.466][c] |

Values correspond to airway smooth muscle mass (α-SMA⁺ airway smooth muscle bundles), and the frequency of α-SMA⁺TUNEL⁺ and α-SMA⁺c-PARP⁺ cells referenced to the biopsy surface area. Data are presented as mean ± standard error of the mean. The 95% confidence intervals of the mean increment (Δ) estimate the size of the effects where differences are statistically significant, or borderline significant as indicated. [a]: versus control subjects; [b]: versus intermittent asthma [c]: versus moderate asthmatics. *: borderline statistical significance.

#### References (Supplemental Data)


E1. Ministerio de la Presidencia. Real Decreto 1201/2005, de 10 de octubre, sobre protección de los animales utilizados para experimentación y otros fines científicos. Boletín Oficial del Estado núm. 252 de 21 de octubre de 2005, pág. 34367. Available at: http://www.boe.es/boe/dias/2005/10/21/pdfs/A34367-34391.pdf.

E2. Proceedings of the ATS workshop on refractory asthma: current understanding, recommendations, and unanswered questions. American Thoracic Society. *Am J Respir Crit Care Med* 2000;162:2341-2351.

E3. Pepe C, Foley S, Shannon J, Lemiere C, Olivenstein R, Ernst P, Ludwig MS, Martin JG, Hamid Q. Differences in airway remodeling between subjects with severe and moderate asthma. *J Allergy Clin Immunol* 2005;116:544-549.

E4. Global Initiative for Asthma. Global Strategy for Asthma Management and Prevention, 2016. Available from: www.ginasthma.org.

E5. Hsia CC, Hyde DM, Ochs M, Weibel ER. An official research policy statement of the American Thoracic Society/European Respiratory Society: standards for quantitative assessment of lung structure. *Am J Respir Crit Care Med* 2010;181:394-418.

E6. Ramos-Barbon D, Presley JF, Hamid QA, Fixman ED, Martin JG. Antigen-specific CD4+ T cells drive airway smooth muscle remodeling in experimental asthma. *J Clin Invest* 2005;115:1580-1589.

E7. Ramos-Barbon D, Fraga-Iriso R, Brienza NS, Montero-Martinez C, Verea-Hernando H, Olivenstein R, Lemiere C, Ernst P, Hamid QA, Martin JG. T Cells localize with proliferating smooth muscle alpha-actin+ cell compartments in asthma. *Am J Respir Crit Care Med* 2010;182:317-324.

E8. Marinas-Pardo L, Mirones I, Amor-Carro O, Fraga-Iriso R, Lema-Costa B, Cubillo I, Rodriguez Milla MA, Garcia-Castro J, Ramos-Barbon D. Mesenchymal stem cells regulate airway contractile tissue remodeling in murine experimental asthma. *Allergy* 2014;69:730-740.
