## Supplementary material for "Contractile cell apoptosis regulates airway smooth muscle remodeling in asthma": ARRIVE Checklist

NOTE: Please save this file locally before filling in the table, DO NOT work on the file within your internet browser as changes will not be saved. Adobe Acrobat Reader (available free here) is recommended for completion.

### The ARRIVE guidelines 2.0: author checklist

#### The ARRIVE Essential 10

These items are the basic minimum to include in a manuscript. Without this information, readers and reviewers cannot assess the reliability of the findings.

| Item | | Recommendation | Section/line number, or reason for not reporting |
|---|---|---|---|
| **Study design** | 1 | For each experiment, provide brief details of study design including: | |
| | | a. The groups being compared, including control groups. If no control group has been used, the rationale should be stated. | Results section with study design graphed in Figs. 1 and 2. |
| | | b. The experimental unit (e.g. a single animal, litter, or cage of animals). | Supplemental Methods section. |
| **Sample size** | 2 | a. Specify the exact number of experimental units allocated to each group, and the total number in each experiment. Also indicate the total number of animals used. | Supplemental data tables and supplememntal Methods section. |
| | | b. Explain how the sample size was decided. Provide details of any *a priori* sample size calculation, if done. | Supplemental Methods section. |
| **Inclusion and exclusion criteria** | 3 | a. Describe any criteria used for including and excluding animals (or experimental units) during the experiment, and data points during the analysis. Specify if these criteria were established *a priori*. If no criteria were set, state this explicitly. | Supplemental Methods section. |
| | | b. For each experimental group, report any animals, experimental units or data points not included in the analysis and explain why. If there were no exclusions, state so. | Supplemental Methods section. |
| | | c. For each analysis, report the exact value of *n* in each experimental group. | Supplemental data tables. |
| **Randomisation** | 4 | a. State whether randomisation was used to allocate experimental units to control and treatment groups. If done, provide the method used to generate the randomisation sequence. | Supplemental Methods section |
| | | b. Describe the strategy used to minimise potential confounders such as the order of treatments and measurements, or animal/cage location. If confounders were not controlled, state this explicitly. | Supplemental Methods section. |
| **Blinding** | 5 | Describe who was aware of the group allocation at the different stages of the experiment (during the allocation, the conduct of the experiment, the outcome assessment, and the data analysis). | Supplemental Methods section. |
| **Outcome measures** | 6 | a. Clearly define all outcome measures assessed (e.g. cell death, molecular markers, or behavioural changes). | Methods, Results, Supplemental Methods and data tables. |
| | | b. For hypothesis-testing studies, specify the primary outcome measure, i.e. the outcome measure that was used to determine the sample size. | Supplemental Methods section. |
| **Statistical methods** | 7 | a. Provide details of the statistical methods used for each analysis, including software used. | Methods section. |
| | | b. Describe any methods used to assess whether the data met the assumptions of the statistical approach, and what was done if the assumptions were not met. | Methods section. |
| **Experimental animals** | 8 | a. Provide species-appropriate details of the animals used, including species, strain and substrain, sex, age or developmental stage, and, if relevant, weight. | Supplemental Methods section. |
| | | b. Provide further relevant information on the provenance of animals, health/immune status, genetic modification status, genotype, and any previous procedures. | Supplemental Methods section. |
| **Experimental procedures** | 9 | For each experimental group, including controls, describe the procedures in enough detail to allow others to replicate them, including: | |
| | | a. What was done, how it was done and what was used. | Methods and Supplemental Methods. |
| | | b. When and how often. | Methods and Supplemental Methods. |
| | | c. Where (including detail of any acclimatisation periods). | Methods and Supplemental Methods. |
| | | d. Why (provide rationale for procedures). | Introduction, Results, Discussion. |
| **Results** | 10 | For each experiment conducted, including independent replications, report: | |
| | | a. Summary/descriptive statistics for each experimental group, with a measure of variability where applicable (e.g. mean and SD, or median and range). | Results and supplemental data tables. |
| | | b. If applicable, the effect size with a confidence interval. | Supplemental data tables. |

### The Recommended Set

These items complement the Essential 10 and add important context to the study. Reporting the items in both sets represents best practice.

| Item | | Recommendation | Section/line number, or reason for not reporting |
|---|---|---|---|
| **Abstract** | 11 | Provide an accurate summary of the research objectives, animal species, strain and sex, key methods, principal findings, and study conclusions. | Abstract (250-word limit) |
| **Background** | 12 | a. Include sufficient scientific background to understand the rationale and context for the study, and explain the experimental approach. | Introduction and Results sections |
| | | b. Explain how the animal species and model used address the scientific objectives and, where appropriate, the relevance to human biology. | Discussion section. |
| **Objectives** | 13 | Clearly describe the research question, research objectives and, where appropriate, specific hypotheses being tested. | Introduction and Results sections. |
| **Ethical statement** | 14 | Provide the name of the ethical review committee or equivalent that has approved the use of animals in this study, and any relevant licence or protocol numbers (if applicable). If ethical approval was not sought or granted, provide a justification. | Methods section. |
| **Housing and husbandry** | 15 | Provide details of housing and husbandry conditions, including any environmental enrichment. | Supplemental Methods section. |
| **Animal care and monitoring** | 16 | a. Describe any interventions or steps taken in the experimental protocols to reduce pain, suffering and distress. | Supplemental Methods section. |
| | | b. Report any expected or unexpected adverse events. | Results section. |
| | | c. Describe the humane endpoints established for the study, the signs that were monitored and the frequency of monitoring. If the study did not have humane endpoints, state this. | Supplemental Methods section and referenced in main Methods. |
| **Interpretation/ scientific implications** | 17 | a. Interpret the results, taking into account the study objectives and hypotheses, current theory and other relevant studies in the literature. | Discussion section. |
| | | b. Comment on the study limitations including potential sources of bias, limitations of the animal model, and imprecision associated with the results. | Supplemental Methods Section and Discussion. |
| **Generalisability/ translation** | 18 | Comment on whether, and how, the findings of this study are likely to generalise to other species or experimental conditions, including any relevance to human biology (where appropriate). | Discussion section. |
| **Protocol registration** | 19 | Provide a statement indicating whether a protocol (including the research question, key design features, and analysis plan) was prepared before the study, and if and where this protocol was registered. | Methods section. |
| **Data access** | 20 | Provide a statement describing if and where study data are available. | Statement in medRxiv upload system. |
| **Declaration of interests** | 21 | a. Declare any potential conflicts of interest, including financial and non-financial. If none exist, this should be stated. | Manuscript title page. |
| | | b. List all funding sources (including grant identifier) and the role of the funder(s) in the design, analysis and reporting of the study. | Manuscript title page. |
